# ‘Finding a Relationship’ Conversations Between Mental Health and Social Care Staff, and Service Users

**DOI:** 10.1101/2024.05.09.24307104

**Authors:** Angelica Emery-Rhowbotham, Helen Killaspy, Sharon Eager, Brynmor Lloyd-Evans

**Affiliations:** Division of Psychiatry, University College London, London, United Kingdom

**Keywords:** Intimacy, Romance, Relationship seeking, Mental illness, Mental health staff, Social care staff, Qualitative, Quantitative

## Abstract

**Background:** Most people seek to establish romantic or intimate relationships in life, including people with mental health problems. However, this has been a neglected topic in mental health practice and research. This study aimed to investigate views of mental health and social care staff about the appropriateness of helping service users with romantic relationships, barriers to doing this, and suggestions for useful ways to support this.

**Methods:** An online survey comprising both closed, multiple response and free-text questions was circulated to mental health organisations across the U.K. via social media, professional networks and use of snowballing sampling. A total of 63 responses were received. Quantitative data were analysed using descriptive statistics, and are reported as frequencies and percentages. Qualitative data were interpreted using thematic analysis, using an inductive approach.

**Results:** Although most participants reported that ‘finding a relationship’ conversations were appropriate in their job role, many barriers to supporting service users were identified, including: a lack of training; concerns about professional boundaries; concerns about service user capacity and vulnerability; and concerns about being intrusive. Participant suggestions for future support included educating service users on safe dating behaviours, and practical interventions such as assisting service users to use dating sites and engage with social activities to develop social skills and meet others.

**Conclusions:** Staff were willing to help service users seek an intimate relationship but may need specific training or guidance to facilitate this confidently and safely. This study elucidates the need for further research in this area, particularly in understanding service user perspectives, and in developing resources to support staff in this work.

## Background

Intimate relationships are a “central aspect of being human” [1] and a “fundamental human right for all” [2]. They affect our environment, quality of life, and without them, basic psychological needs remain unfulfilled [3]. Intimate relationships, as discussed in this work, encompass those distinct from platonic friendships in involving physical intimacy, sexual activity, or romantic love [4]. They may include both monogamous and non-monogamous relationships.

Social relationships are associated with many facets of psychological health, including feelings of self worth and self-esteem and low levels of depression, anxiety, and substance use [5–7]. The quality and satisfaction of an intimate relationship may have a particularly important impact on wellbeing due to potentially heightened positive emotions and cognitions [8]. For those with a mental health condition, evidence shows various benefits of an intimate relationship, including: providing companionship [9]; helping individuals to stay calm and relaxed [10]; providing emotional support [11]; instilling confidence [12]; and allowing sexual and emotional expression [13].

Moreover, loneliness is a common problem reported amongst individuals with mental health problems, which may increase one’s risk of both physical and mental health problems, and predict poor recovery for those with an existing mental health problem [14]. For instance, up to 40% of individuals with depression report feeling lonely most of the time [15] and their odds of feeling lonely are ten times that of the general population [16]. Loneliness has been conceptualised as having three dimensions: intimate, relational, and collective [17]. While the latter two indicate a lack of wider social networks such as friends and communities, intimate loneliness indicates a lack of a close emotional attachment, and not being able to share intimacy with another [18]. While having an intimate partner or spouse can reduce one’s level of intimate loneliness [19–21], loneliness interventions rarely, if ever, address an individual’s need or desire for an intimate relationship [22]. Thus, investigation into whether and how mental healthcare services could support people to achieve desired intimate relationships, and combat intimate loneliness, is warranted.

The most recent review of mental health service users’ views on sexuality was published by McCann and colleagues in 2019 [3], which found that sexuality is often a neglected topic both by mental health practitioners and service users. Most studies discussing intimacy and mental illness, represented in this review and in the wider literature, are focused on sexuality in populations with severe mental illness, and consistently find that mental health staff typically do not discuss people’s needs and wishes for intimacy and romantic relationships [23,24]. Furthermore, this limited research primarily focuses on sexual health and diminishing the risk of sexually transmitted diseases, and neglects the positive rewards of intimate relationships such as sexual pleasure, connection and commitment [25–27].

The limited literature that has focused on the wider experience of intimate relationships suggests that mental health service users may benefit from support to achieve such a relationship. Besides struggling with maintaining relationships - for reasons including chronic low relationship satisfaction [28], and a hesitancy to trust and be intimate with another [29] - those with mental illness consistently report struggling to establish an intimate relationship in the first place. Relatively few people with serious mental illness (15% in one study [30]) have romantic relationships when compared to the general population [31,3], despite 71% spontaneously identifying relationships, intimacy and sexuality as facilitating recovery [30]. The gap between wanting a relationship, but not attaining one, may be partly explained by stigma [32,30]. Societally, those with mental illness are often heavily stigmatised, for instance in being rated as below average on several factors related to mate selection, such as social status, sexual desirability and personality [33–35]. Furthermore, those with mental illness are often aware of these stigmatising attitudes, and can even develop self- stigmatising attitudes also [36]. For instance, 66-89% of psychiatric outpatients agreed that most people ‘do not have an interest in having a romantic / sexual relationship with someone who has a mental illness’ [37]. This leads to active avoidance of intimate relationships [32], contributing to the finding that, globally, 40% of those with depression have intentionally avoided initiating a close relationship [38].

In addition to stigma, reasons for not seeking an intimate relationship include the direct effects of symptoms such as poor self-esteem and low motivation; medication side effects such as extinguished libido [39], and lack of opportunity to meet potential partners, particularly while being an inpatient [30,40]. It is also important to note the potential damaging effect of intimate relationships for those with mental illness. For instance, existing mental health problems can increase one’s vulnerability to intimate partner violence [41], and relationship loss can have ‘devastating’, effects of further social losses, increased loneliness, and a regression in one’s recovery [21,22]. Embarking on an intimate relationship should therefore be done under careful consideration by both the service user and their mental healthcare provider.

Despite these key obstacles however, service users recognise both “being lonely” and a “lack of a significant relationship” as being direct barriers to their recovery [30]. And, when asked about this directly, many service users express a desire for help in attaining an intimate relationship [40], despite this being rarely offered [42].

In recent years, the adoption of recovery-oriented approaches in mental health care has increased focus on empowering people to achieve a meaningful, fulfilling life alongside their mental health problems, rather than symptom eradication being the main focus [43]. This includes helping people to build relationships, which is explicitly included in policy in many health care systems. In the U.K. for example, The Care Act 2014 specifies developing and maintaining personal relationships as an eligible need for support [44].

Despite this, relationships of an intimate nature are not routinely discussed in clinical mental health settings, and have not often been the focus of research [29,37]. Limited literature that investigates this omission suggests that mental healthcare staff are ambivalent as to whether they should support service users in the domain of intimate relationships. While staff note that having such a relationship can aid the progression of treatment [45], they also reference several barriers to discussing this subject with service users. These include: a lack of professional training or skills; uncertainty about the appropriateness of such conversations; personal discomfort around discussing sexuality; and organisational factors such as a lack of time and resources [23,46].

Due to these issues, intimate relationships tend to have low priority within mental healthcare [47,48] despite service users’ willingness and the recognition of its relevance to therapeutic goals [3,49]. However, unless their clinician brings it up, service users tend to feel that conversations about intimacy are ‘out of bounds’ [49], except when relationships are abusive or potentially contributing to their illness [42,50]. Equally, mental health practitioners often believe that service users should lead on raising the subject of intimacy [51,45]. For instance, over half of clinical psychologists describe “never” or “rarely” discussing issues of intimacy or sexuality [52]. As reported by service users, this can result in the belief that the experience of intimacy is unattainable for them [53].

Little is known about how mental health services could best support service users seeking an intimate relationship. ‘Dating skills’ groups have been trialled and found acceptable [54,55], yet so far these have been limited to male participants with psychosis. There are several additional practical suggestions in the literature, regarding how mental health staff might help service users who are seeking a relationship, including: accompanying service users to social events where they can gain social skills and meet people [32,54]; coaching service users on the use of dating websites and apps [32]; as well as providing education to service users about relationships, and opportunities for peer support [56]. However, none of these examples have been widely implemented or evaluated.

## Rationale, aim and objectives of this study

Given the importance of intimate relationships in people’s lives; the specific barriers for people with mental health problems in finding relationships; and the general lack of knowledge about how to support service users in this domain, this study aimed to investigate: the perspectives of U.K. mental health and social care staff on supporting service users to find an intimate relationship; barriers to doing so; and suggestions as to how to increase support to enable this.

## Methods

### Research questions

Due to the limited existing literature on this topic, the current study took an exploratory approach to address three research questions:

1. What are staff perspectives around the appropriateness of supporting service users with achieving desired intimate relationships?
2. What helps and hinders staff to have ‘finding a relationship’ conversations?
3. What strategies can staff use to support people with finding a relationship?

### Design

This study employed a cross-sectional, mixed-methods design, to collect both quantitative and qualitative data through an online survey. Quantitative data were collected using closed, multiple choice survey questions, and qualitative data were collected using open, free text response questions. The survey was disseminated to mental health and social care staff within the U.K. using snowballing sampling. This study was approved by the UCL Research Ethics Committee on the 7^th^ March 2023 (Project ID: 24833/001).

### Setting

This study comprised an online survey constructed using Qualtrics software [version XM, 2023]. Respondents were directed to a link to the survey which took around 15 minutes to complete.

### Participants

Mental health staff from any professional group were eligible to take part. This included clinically trained staff such as psychiatrists, psychologist nurses, social workers and occupational therapists, as well as non-clinically qualified staff such as support workers, peer workers and social care staff. Participants were excluded if they worked in a specialist relationship counselling, sexuality or gender identity service as the study wished to focus on general mental health and social care services.

### Materials

The online survey questionnaire began by asking the participant to provide demographic information including their age, gender and ethnicity, as well as information about how long and in what capacity they had worked in mental health services. The survey questionnaire then asked whether the participant helped service users to find a relationship and if so, how; any barriers and facilitators they experienced in having ‘finding a relationship’ conversations with service users; any training and other support they had received in this area; as well as their opinions on particular methods of helping service users to find relationships. The matrices for rating particular barriers, and methods of helping service users, were created referencing staff responses in a recent focus group study [40]. The full survey questionnaire is provided in Appendix A.

### Procedures

The survey was open for 11 weeks (15^th^ May to 31^st^ July 2023), dictated by a UCL Master’s programme deadline. All participants provided their informed consent online, before proceeding to the survey questions (see Appendix A for the study information provided in the online survey homepage). For recruitment, invitation emails were sent to a list of professional contacts, compiled using the professional networks of each member of the research team. This included national networks for social workers, psychiatrists and nurses, and national voluntary sector organisations providing mental health care. Invitations were extended to both individuals and organisational contacts, and each contact was asked to further disseminate the survey in turn, in a snowballing approach. Social media outreach included posting on the organisational X (formerly Twitter) accounts of the research team, and sharing on mental health spaces on Facebook, LinkedIn and Reddit.

Once participants had clicked on the invitation link to the survey, they first read through all participant information and data protection information and, if satisfied, then clicked a check box to confirm their consent. They then proceeded to complete the questionnaire on Qualtrics [version XM, 2023]. Before finishing, participants were given the option to provide their email address if they wished to receive a final report of the study, or to agree to be contacted about participation in future, related studies.

While recruitment was ongoing, data was stored within the Qualtrics programme, and once data collection had ended, the survey was deleted from Qualtrics and all survey data were moved to UCL’s secure online folders.

### Analysis

#### Quantitative

For the 12 closed, multiple choice questions, descriptive statistics were analysed using *Microsoft Excel.* Participant demographic and job-related characteristics, as well as other quantitative, opinion-based questions were reported using frequencies and percentages.

#### Qualitative

For the free text responses, Braun and Clarke’s [60] thematic analysis approach was utilised. An inductive approach to thematic analysis was used, meaning analysis of themes and sub- themes was data-driven and not informed by a theoretical framework [61]. There were seven distinct free text response questions from the survey. One question asked about perceived appropriateness of ‘finding a relationship’ support, four asked about barriers to offering this support, and two asked about methods and suggestions to increase such support. Analysis was led by AER and undertaken in six phases. First, data familiarisation involved reading over all participant responses. Second, these responses were coded into ‘meaningful’ units of text. The third phase entailed organising these codes into themes i.e. codes which could fall under the same category were grouped together and given a label. Phases four and five involved reviewing and agreeing the labelling of themes and sub-themes through discussion with the research team, and refining labels by consensus where appropriate. It was at this point that the seven original questionnaire items were merged into three themes, (appropriateness, barriers, and methods/suggestions). The final phase involved producing an analytical report (for a full breakdown of the analysis process, see Appendix B).

### Reflexive statement

A researcher’s personal characteristics and positioning inevitably shapes their understanding and analysis of material, particularly within thematic analysis due to the heightened subjectivity of researcher-led generation of themes [57]. Thus, acknowledging one’s positioning using reflexive practice is vital as, being privy to the assumptions underlying the analysis, both author and reader are able to challenge them, protecting the work from undue bias [58]. In the current study, all co-authors are white, three are female, and all are educated to at least post-graduate level. The lead author has little experience of working with adults in a mental healthcare role. One co-author is a senior clinical academic psychiatrist and thus has relevant clinical experience, limiting misinterpretation of findings. Other co-authors include an early career researcher and a senior mental health academic with a background in social work. To reduce subjectivity bias, a coding diary was kept, which was used to reflect on participant responses which elicited an emotional response, or were felt to be puzzling. These reflections were discussed with the research team at regular intervals and informed the interpretation of the data.

## Results

### Participant characteristics: quantitative results

A total of 63 mental health and social care staff participated in the survey, and 44 completed all questions. The majority of respondents were female (n=54), most were white (n=49), in the age range 26-35 (n=24), and did not follow a particular religion (n=44). The number of years participants had worked in mental health services was relatively evenly split, with a small majority having worked for 2-5 years (n=19). Most reported their profession to be psychologists (n=16), who were working in the NHS (n=46) in a community based mental health team (n=32). See Table 1 for a full breakdown of participant characteristics, and see Appendix C for all quantitative results tables. (Note: Tables which are longer than an A4 page are placed at the end of the document).

**Table 1.** Participant characteristics

| **Variables**<br>**(Total N responses)** |  | **n (%)** |
| --- | --- | --- |
| Gender<br>(N = 63) | Female | 54 (85.7) |
|  | Male | 9 (14.3) |
| Age<br>(N = 63) | 18-25 | 12 (19.1) |
|  | 26-35 | 24 (38.1) |
|  | 36-45 | 15 (23.8) |
|  | 46-55 | 8 (12.7) |
|  | 56-65 | 4 (6.4) |
|  | 65+ | 0 (0) |
| Ethnicity<br>(N = 63) | White | 49 (77.8) |
|  | Asian or Asian British | 6 (9.5) |
|  | Mixed or multiple ethnic groups | 5 (7.9) |
|  | Black, black British, Caribbean or African | 2 (3.2) |
| Religion<br>(N = 63) | No religion | 44 (69.8) |
|  | Christianity | 10 (15.9) |
|  | Buddhism | 3 (4.8) |
|  | Islam | 3 (4.8) |
|  | Hinduism | 1 (1.6) |
|  | Other | 1 (1.6) |
|  | Judaism | 0 (0) |
|  | Sikhism | 0 (0) |
| Years in mental health<br>services<br>(N = 60) | Less than 2 years | 11 (18.3) |
|  | 2-5 years | 19 (31.7) |
|  | 6-10 years | 14 (23.3) |
|  | More than 10 years | 16 (26.7) |
| Professional group<br>(N = 60) | Psychologist | 16 (26.7) |
|  | Occupational therapist | 9 (15.0) |
|  | Psychiatrist | 9 (15.0) |
|  | Support worker | 9 (15.0) |
|  | Nurse | 6 (10.0) |
|  | Peer support worker | 3 (5.0) |
|  | Social worker | 3 (5.0) |
|  | Counsellor / therapist | 2 (3.3) |
|  | Other | 2 (3.3) |
| Sector<br>(*N* = 60) | NHS | 46 (76.7) |
|  | Independent sector | 7 (11.7) |
|  | Voluntary sector | 5 (8.3) |
|  | Local authority | 0 (0) |
| Service type<br>(*N* = 60) | NHS community mental health team | 33 (55.0) |
|  | Supported accommodation | 10 (16.7) |
|  | Inpatient | 9 (15) |
|  | Other | 5 (8.5) |

### Appropriateness of finding a relationship conversations: quantitative results

Participants rated how far they agreed that ‘finding a relationship’ conversations were appropriate in their work role; 70% reported that they either ‘strongly’ or ‘somewhat’ agreed, and 8% ‘strongly’ disagreed (see Table 2).

**Table 2.** Ratings of agreement as to the appropriateness of ‘finding a relationship’ conversations

| Agreement as to appropriateness of ‘finding a relationship’ conversations | *n* (%)<br>(total N = 50) |
| --- | --- |
| Strongly agree | 10 (20.0) |
| Somewhat agree | 25 (50.0) |
| Somewhat disagree | 11 (22.0) |
| Strongly disagree | 4 (8.0) |

### Appropriateness of finding a relationship conversations: qualitative results

A total of 45 participants responded to the free-text question of why they felt providing ‘finding a relationship’ support was, or was not, appropriate in their job role (see Table 3 below).

**Table 3.** Appropriateness of ‘finding a relationship’ support: themes, sub-themes, example quotes and number of contributing participants

| Theme | Illustrative quote [participant ID] | Number of participants who contributed to theme (total N = 45) |
| --- | --- | --- |
| *Subtheme* |  |  |
| **Benefits to having ‘finding a relationship’ conversations** |  | **35/45** |
| *Encouraging recovery* | “I feel that romantic relationships and intimacy are a human need and have a huge impact on mental health” [ppt. 36] | 15 |
| *Relevance to service users* | “I believe this is important. Because many service users can struggle to form relationships in general, and having some support regarding intimate relationship would be useful for these users” [ppt. 58]. | 11 |
| *Methods to support service user relationships* | “We should support our service users to recognise healthy relationships, build boundaries and encourage positive social interactions [ppt. 9]” | 6 |
| *Having safe boundaries and limits* | “I think its only our work role if its [mental health] related” [ppt. 29] | 5 |
| **Reasons to not have ‘finding a relationship’ conversations** |  | **13/45** |
| *Moral and ethical issues* | “I feel like it would be unethical to help them find a relationship” [ppt. 3] | 8 |
| *Not feasible in my job role* | “Actually practically finding somebody feels a little beyond our scope” [ppt. 35] | 5 |

Overall, 35 respondents expressed opinions about the benefits of having ‘finding a relationship’ conversations and for the most part, these were felt to be helpful in encouraging recovery. For some, this was mentioned in relation to a holistic model of practice, noting that staff should be working with “all aspects of life” [ppt. 48]. Otherwise, participants made an explicit link between having an intimate relationship and being better able to manage one’s mental health and loneliness.

Secondly, participants discussed how ‘finding a relationship’ conversations were generally relevant to service users. This was expressed by some in terms of particular service users, as they may “struggle with intimacy” [ppt. 26], or with “form[ing] relationships in general” [ppt. 58], rendering relationship seeking a relevant therapeutic goal. Others reflected that relationship attainment is desired by service users, and thus it is appropriate to “help/support/advise in ways I can” [ppt. 24].

Participants also reflected on specific methods to support service user relationships, and how these are particularly appropriate to engage in. Discussed here was the importance of supporting service users to recognise what is safe and acceptable in a relationship, as well as self-esteem building, skills building, and increasing social opportunities. Finally, some participants expressed the need for safe boundaries and limits within ‘finding a relationship’ conversations. For instance, one participant expressed that this work is appropriate “as long as you keep it within certain boundaries” [ppt. 33], with another expressing: “I think it’s only our work role if it’s [mental health] related, not just because someone without [mental health] can’t find a relationship” [ppt. 41]. (Note: both of these topics were discussed more completely later in the survey – see the ‘barriers’ and ‘methods and suggestions for support’ sections below for further discussion).

Fewer participants (13/45) expressed reasons not to engage in ‘finding a relationship’ conversations. Of those who did, eight discussed moral and ethical issues. For instance, as one stated directly: “I feel like it would be unethical to help them find a relationship” [ppt. 3].

Participants also reported ‘finding a relationship’ conversations feeling inappropriate due to their intrusiveness, or because of risks including service users’ potential for exploitation, history of violence towards others, or due to concerns about professional culpability if something went wrong.

Some participants felt that ‘finding a relationship’ conversations were not feasible in their job role. This was for varying reasons, such as having a highly specified job description, where new goals could not be easily added to their agenda. Others noted a “lack of funding and commissioning” which limits staff to only offering interventions with clear “mental-health related outcomes” [ppt. 51]. Others discussed relying on other professionals such as “support workers” [ppt. 38] or “social workers / occupational therapists” [ppt. 43], roles in which they believed ‘finding a relationship’ conversations may be more appropriate.

### Barriers to helping service users find a relationship: quantitative data

Overall, participants had mixed opinions about which of the specified barriers were the most important. Rated the most important (highest rating for ‘a great deal’ of importance) was lack of training, followed by worries about professional boundaries, service user vulnerability, and intrusiveness. Some barriers were rated consistently as ‘not at all important’ by the majority, such as lack of management support, lack of time, and lack of training.

Lack of training and worries about professional boundaries were selected by some participants as highly important barriers, but were rated as unimportant barriers by other participants (see Table 4 below for a full outline of responses).

**Table 4.**
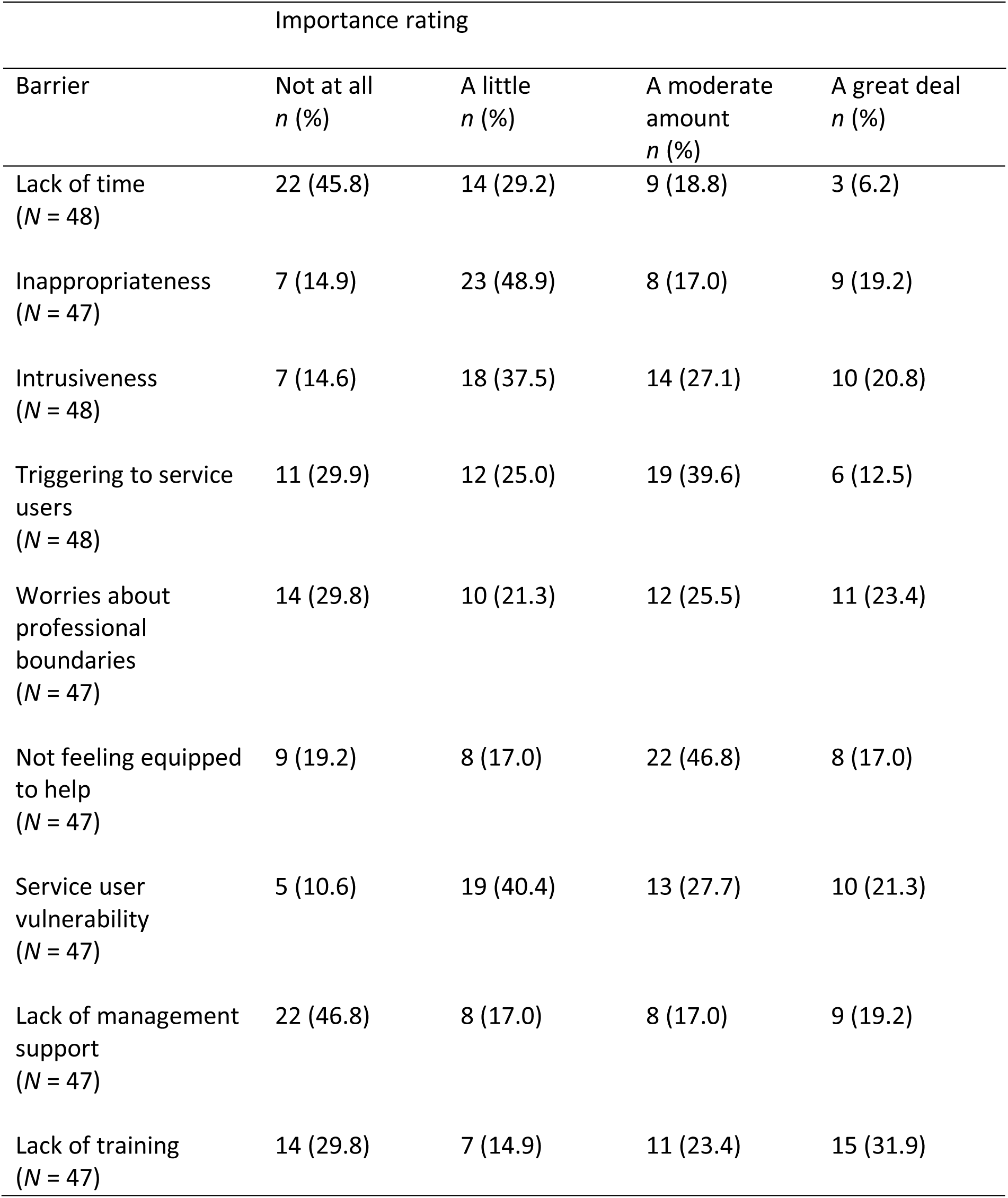
Participant ratings of the importance of potential barriers to helping service users find intimate relationships.

### Barriers to helping service users find a relationship: qualitative data

Across three free text response questions, participants were asked to elaborate on any barriers that they perceived as being discouraging of ‘finding a relationship’ conversations. These are summarised in Table 5.

**Table 5.**
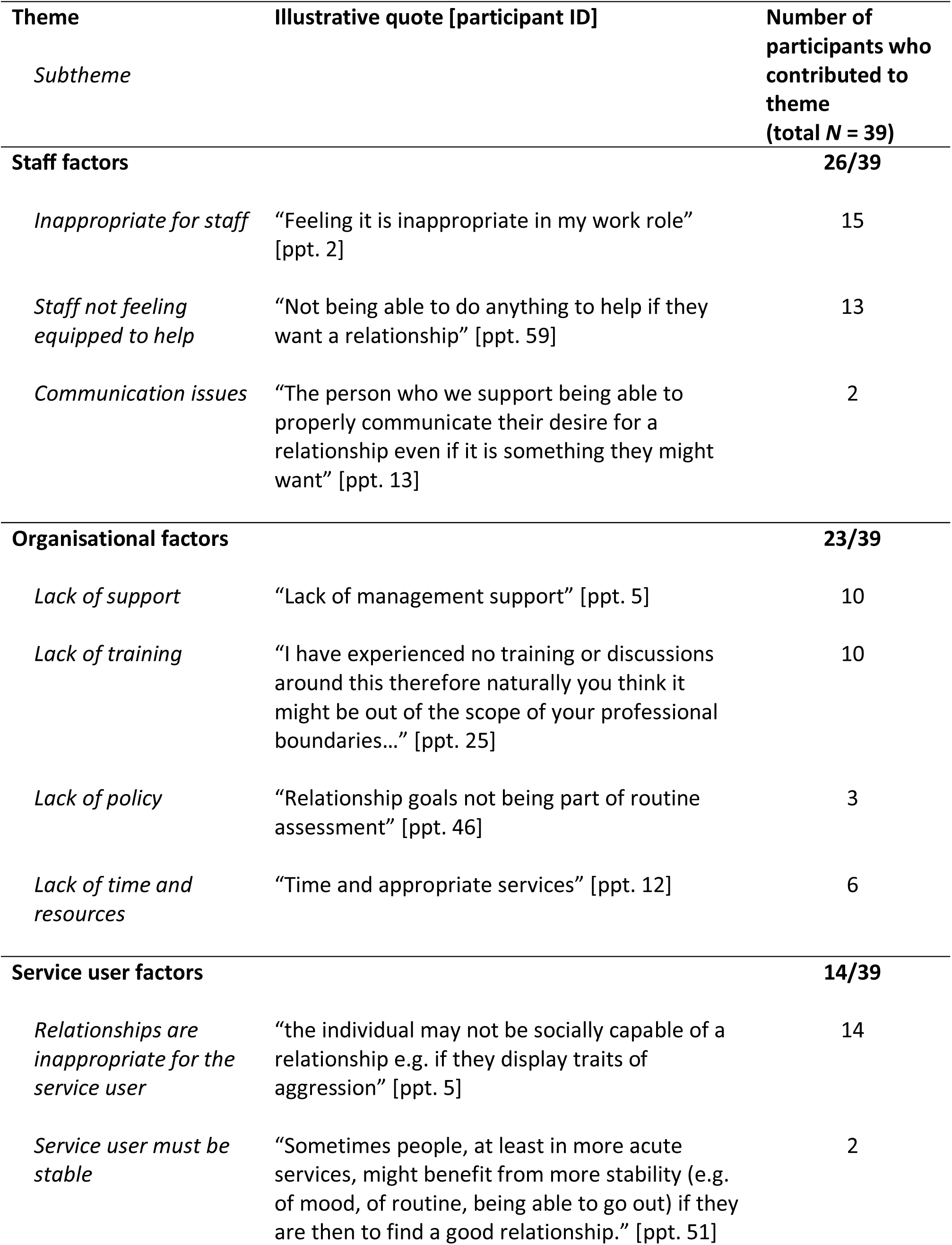
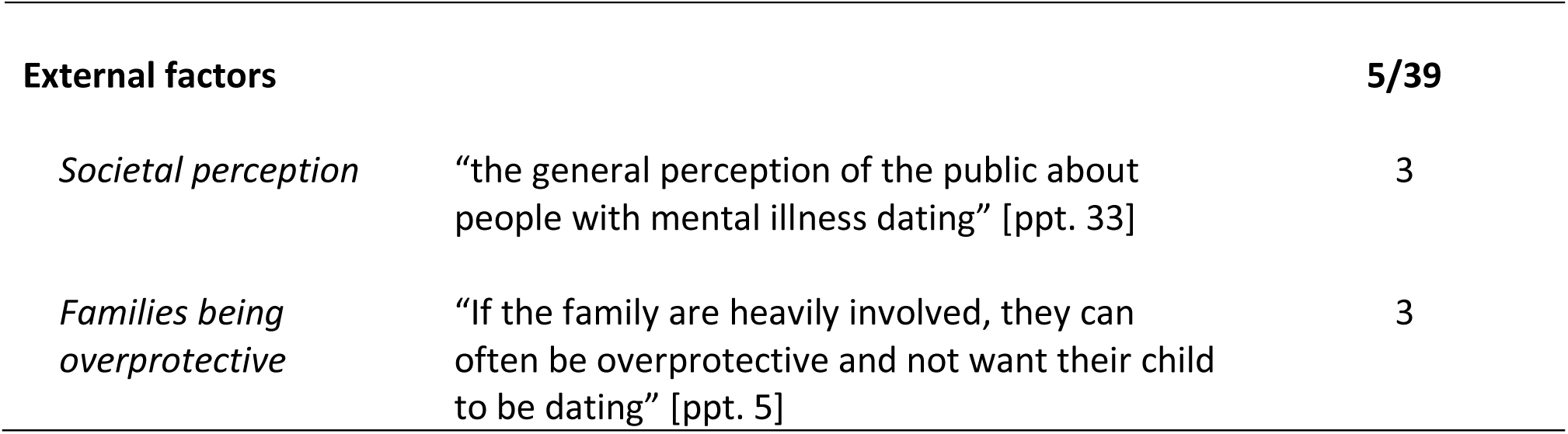
Barriers to finding a relationship support: themes, sub-themes, examples and number of contributing participants

| Theme | Illustrative quote [participant ID] | Number of participants who contributed to theme (total N = 39) |
| --- | --- | --- |
| *Subtheme* |  |  |
| **Staff factors** |  | **26/39** |
| *Inappropriate for staff* | "Feeling it is inappropriate in my work role" [ppt. 2] | 15 |
| *Staff not feeling equipped to help* | "Not being able to do anything to help if they want a relationship" [ppt. 59] | 13 |
| *Communication issues* | "The person who we support being able to properly communicate their desire for a relationship even if it is something they might want" [ppt. 13] | 2 |
| **Organisational factors** |  | **23/39** |
| *Lack of support* | "Lack of management support" [ppt. 5] | 10 |
| *Lack of training* | "I have experienced no training or discussions around this therefore naturally you think it might be out of the scope of your professional boundaries..." [ppt. 25] | 10 |
| *Lack of policy* | "Relationship goals not being part of routine assessment" [ppt. 46] | 3 |
| *Lack of time and resources* | "Time and appropriate services" [ppt. 12] | 6 |
| **Service user factors** |  | **14/39** |
| *Relationships are inappropriate for the service user* | "the individual may not be socially capable of a relationship e.g. if they display traits of aggression" [ppt. 5] | 14 |
| *Service user must be stable* | "Sometimes people, at least in more acute services, might benefit from more stability (e.g. of mood, of routine, being able to go out) if they are then to find a good relationship." [ppt. 51] | 2 |
| **External factors** |  | **5/39** |
| *Societal perception* | “the general perception of the public about people with mental illness dating” [ppt. 33] | 3 |
| *Families being overprotective* | “If the family are heavily involved, they can often be overprotective and not want their child to be dating” [ppt. 5] | 3 |

In total, 23 of 39 respondents discussed staff factors as being notable barriers to ‘finding a relationship’ conversations, of whom 15 expressed ideas surrounding the perceived inappropriateness of relationship seeking support, especially due to feelings of intrusiveness and breaking professional boundaries. In addition, 10 respondents discussed not feeling equipped to help. Often this was due to fears about making things worse, for example “stigmatis[ing] [service users’] single status” [ppt. 40], or “com[ing] across as condescending” [ppt. 13]. Some participants expressed “not feeling able to help” [ppt. 57] or else having “low confidence” [ppt. 24] in matters related to relationships.

Approximately half (23) of the respondents also discussed organisational factors as a notable barrier. Ten described a lack of support, as one participant shared: “everything you do has to be mostly approved or encouraged by them, so … without management support, it is not something that can be done” [ppt. 5]. Ten participants reflected on the need for more staff training, as having “experienced no training or discussions around this … naturally you think it might be out of the scope of your professional boundaries” [ppt. 31]. Otherwise, six participants reported not having enough time or resources to focus on relationship conversations. As one participant said: “there is barely enough time to do the core aspects of my job, so … there is very unlikely to be resource for this” [ppt. 43].

Service user factors were cited by 14 of 39 respondents who answered this item. Here, participants expressed ideas regarding the inappropriateness of a relationship for some service users. Some worried about service users, being “vulnerable to exploitation” [ppt. 62], perhaps due to a history of “domestic violence” [ppt. 7], or “sexual assault” [ppt. 41].

Other participants perceived having a relationship would be detrimental to the service user, as it may “present another stressor…” [ppt. 2]. Moreover, some participants mentioned that a service user should be relatively stable before beginning to seek a relationship, as they may be more successful that way.

Finally, five participants mentioned external barriers. This included the families of service users being “overprotective” [ppt. 13], and not wanting them to engage in a relationship.

Participants also mentioned societal pressures and perceptions as being barriers - either that people with mental illness should not be dating, or that staff should not perpetuate the societal pressure that everyone need be in a relationship to be happy.

### Nature of ‘finding a relationship’ conversations with service users: quantitative data

The online survey questions asked participants to report on various aspects of their current practice regarding ‘finding a relationship’ conversations, including their perception of service user interest in finding a relationship. Overall, 64% reported that the majority of their service users were single, and 60% reported that the majority of their service users would not want to find a relationship.

Regarding ‘finding a relationship’ conversations specifically, 64% of respondents reported that they had had conversations with ‘few’ of their service users about finding a relationship, while a small proportion (10%) reported never having done so. Very few respondents reported ‘usually’ broaching the conversation (4%) and none said they ‘always’ brought the subject up. It was reported by most as easier to have ‘finding a relationship’ conversations in one-to-one settings compared to group situations, and finally only 7% reported having received any training on the topic of offering relationship support to service users. (see Table 6 for a full breakdown of these findings).

**Table 6.**
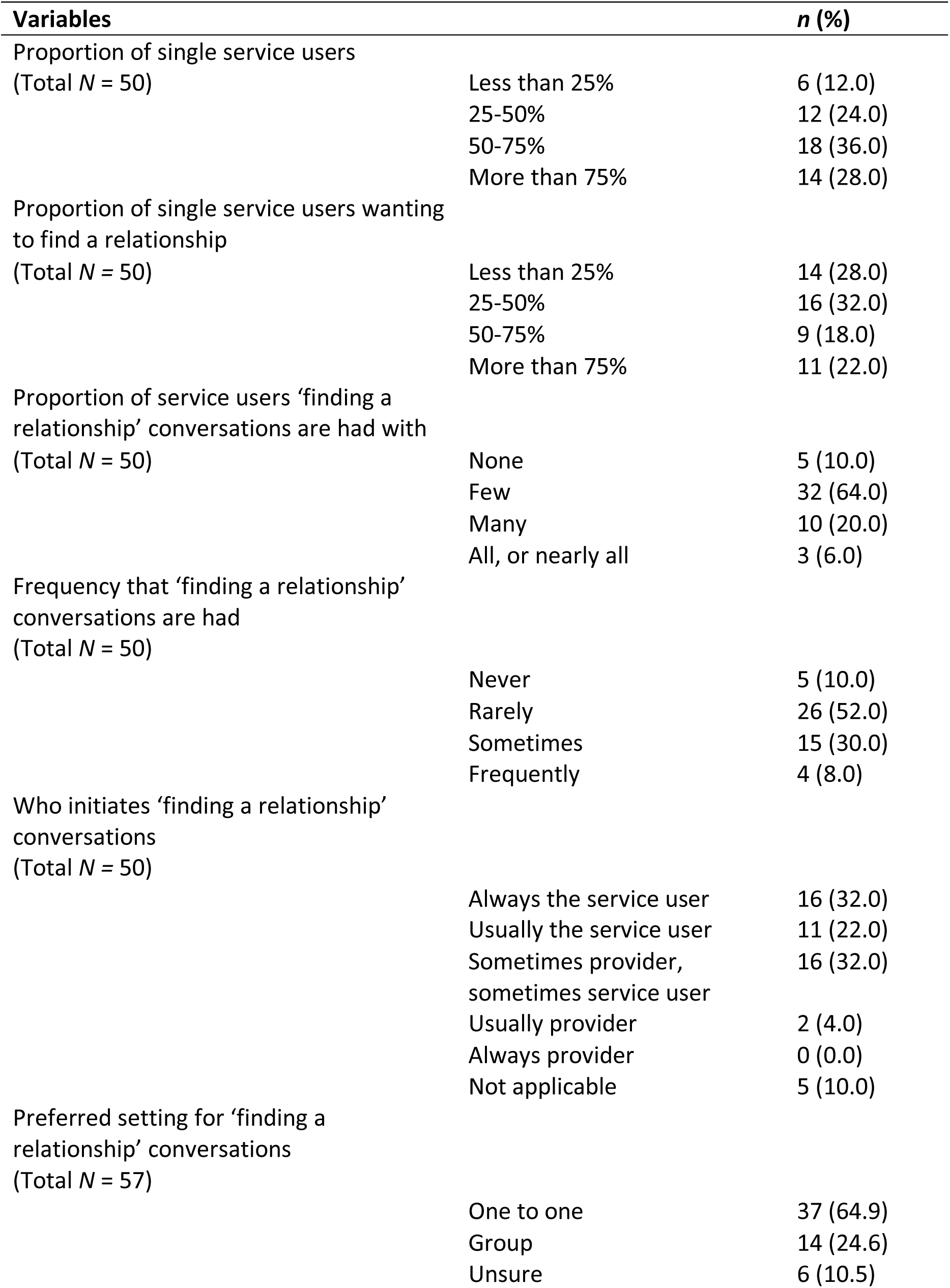
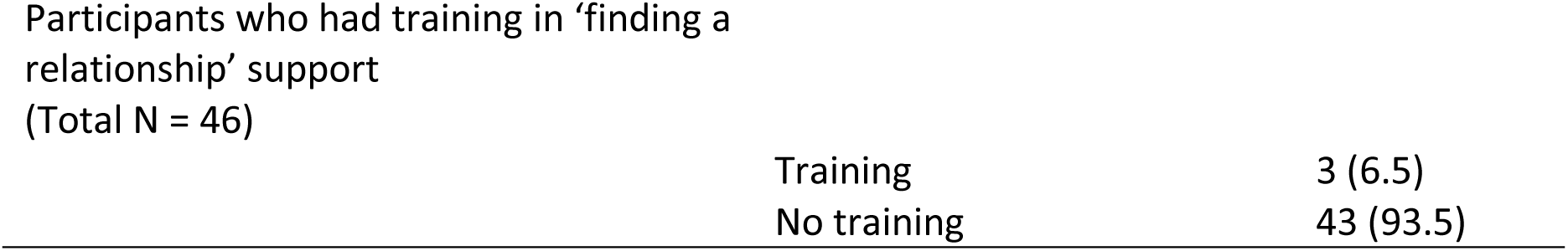
Quantitative findings regarding the nature of ‘finding a relationship’ conversations

### Support and suggestions offered to help service users find a relationship: qualitative data

Participants were asked to suggest any methods that they had used to help service users with relationship seeking, as well as suggestions for methods to use in future practice. This item was answered by 37 respondents, and responses are summarised in Table 7 below.

**Table 7.** ‘Methods of Support’ themes, sub-themes, example quotes and number of contributing participants

| Theme | Illustrative quote [participant ID] | Number of participants responses coded by theme and sub-theme (total N = 37) |
| --- | --- | --- |
| *Subtheme* |  |  |
| **Current practice** |  | **33/37** |
| *Preparing the service user for the dating world* | "We try and sensitively explain how consensual relationships work. Remind them how coercive behaviour is wrong and try and guide them on acceptable behaviour." [ppt. 7] | 15 |
| *Discussions with service user about relationships* | "Just having a conversation about it and finding out their thoughts." [ppt. 33] | 14 |
| *Increasing access to partners* | "Behavioural experiments using dating apps and broadening social opportunities." [ppt. 25] | 5 |
| **Suggestions for future practice** |  | **25/37** |
| *Education and skills work* | "Perhaps psycho[education] on healthy relationships, [and] for some people like LD services maybe learning social skills to develop relationships" [ppt. 47]. | 8 |
| *Increasing access to partners* | "Social opportunities in services" [ppt. 11] | 7 |
| *Systemic change* | "Improving policies and access to material to meet needs" [ppt. 48] | 6 |
| *Open discussion about relationships in service* | "Open conversation. Looking at roles and routines that might support opportunities. We can explore what the emotional, practical and esteem barriers are." [ppt. 49] | 5 |
| *Signposting* | "Direct them to relevant services" [ppt. 8] | 3 |

Regarding current practice, 15 of 37 respondents described preparing the service user for the dating world. This method included educating the service user about dating safely, for instance, one shared that they: “provide support and encouragement, help people to identify their needs and goals, [and] teach people about communication and relationship skills” [ppt. 39]. Some respondents also described helping service users to build skills relevant to dating, such as: “building interpersonal social skills and recognising how they might be vulnerable to exploitation” [ppt. 10], or “building up self-esteem” [ppt. 11].

A total of 14 respondents shared thoughts about their discussions with service users. Discussion types were varied, for instance, some participants described “just having a conversation … and finding out their thoughts” [ppt. 33], while others had more directed conversations, for instance “to ask whether having a relationship is one of their goals” [ppt. 40]. Other discussions involved taking the service user’s lead, and discussing social connections, barriers, and sexual needs.

Increasing access to partners was discussed by five of 37 respondents. This involved directly supporting service users to use dating sites and apps, as well as helping to identify appropriate social opportunities.

Finally, four participants stated they were either “not sure” [ppt. 32] of current practice in their service, or that there was no relationship support provided: “In 20 years I’ve not witnessed this” [ppt. 34].

Suggestions for future practice generally reflected participants’ current practice. For instance, the most cited suggestion was education and skills work, which is in line with preparing the service user for the dating world, discussed above. Here, participants suggested increasing access to group support as a method to discuss “online dating safety” [ppt. 43], or to work through “scenarios [to] explore what they would do in [certain] situation[s]” [ppt. 30]. Some participants also recommended increasing the social skills requisite for developing an intimate relationship.

Seven of the 37 suggested increasing access to partners. For most, this meant helping service users to engage with social activities outside the service e.g. “group activities in the community” [ppt. 4]. Others suggested “social opportunities in services” [ppt. 11], as well as “find[ing] dating services” [ppt. 31] either in person or online.

Respondents also suggested increasing discussion of relationships in the service (5/37). It was highlighted that such discussions be “more open… and not stigmatising [ppt. 33]”, and in such a way as “to give permission to patients to state this as a goal” [ppt. 40].

Finally, six respondents suggested systemic change, noting that mental health organisations need to be “improving policies and access to material to meet needs” [ppt. 48]. Three reported that signposting service users to other services is most appropriate.

## Discussion

The present study made several unique contributions to the topic of relationship seeking support in mental healthcare. Firstly, despite having had ‘few’ and ‘rare’ conversations about romance and intimacy with service users, the current participants mostly reported that ‘finding a relationship’ conversations were appropriate in their job role. Quantitative analysis showed that barriers to ‘finding a relationship’ conversations rated the most important were: a lack of training; concerns about professional boundaries; concerns about service user capacity and vulnerability; and concerns about being intrusive. Respondents reported having engaged in relationship seeking support through discussions with service users, and by taking steps to prepare them for entering the dating world. Participant suggestions for future support included educating service users on safe dating behaviours, and practical interventions such as assisting service users to use dating sites, and engage with social activities to develop social skills and meet others. This is one of very few studies to investigate mental health staff perspectives around the practice of helping service users to seek a relationship [e.g. 59].

Amongst staff respondents, 70% agreed that ‘finding a relationship’ conversations were appropriate in their job role. One recent investigation of mental health practitioner attitudes found that only around 30% of participants expressed that dating and romantic relationships would be legitimate therapeutic goals [23]. These discordant results may be explained by cultural differences between the study settings (U.K. and Israel) and signal the need for more investigation on this topic.

The barriers rated the most important were a lack of training, worries about professional boundaries, and worries about being intrusive. These findings suggest that respondents felt uncomfortable discussing issues related to personal relationships with service users. The concept of ‘boundaries’ in mental healthcare is nuanced and ‘crossing boundaries’ can be any combination of beneficial, neutral or harmful to the service user [62]; however some argue that the inflexible maintenance of professional boundaries perpetuates the power imbalance between service user and practitioner, and can be dehumanising to the former [63]. A series of nine reflective questions have thus been proposed to aid mental health practitioners considering a boundary crossing [64]. One of these is to imagine the ‘best possible outcome’ of crossing the boundary, and the ‘worst possible outcome’ both of crossing, and not crossing the boundary [64]. If relationship seeking support was promoted in mental healthcare, therefore, practitioners may be encouraged to reflect on and re-examine their professional boundaries in relation to this.

Organisational factors were often rated to be unimportant barriers by the staff respondents. These findings diverge from existing literature. For instance, it is reported that a broad lack of guidance leaves mental healthcare staff with a perceived lack of competency, in turn leading to confusion, frustration and even low professional self-esteem [23]. A ‘lack of training’, however, was rated as being ‘not at all’ important and ‘very much’ important by a similar number of participants in the current study. These diverging views may reflect the difference in attitudes within the current sample. For the 30% of participants who disagree that ‘finding a relationship’ conversations are appropriate, they are likely to disagree that a lack of support or training is the key barrier to this, and agree that issues of appropriateness or professional boundaries are the key barrier. Future investigation, therefore, may benefit from stratifying analysis by participants who believe relationship seeking support is appropriate, and those who do not, such that nuances in perceptions may be explored.

Finally, service user factors were commonly discussed, where perceptions centred around service users either being too vulnerable, too volatile, or being in a context which made finding a relationship redundant. In the wider literature, attitudes that romantic and sexual relationships are either wholly ‘irrelevant’ or ‘detrimental’ to service users are discussed as paternalistic and potentially harmful to service user recovery [23]. The literature also cites that healthcare practitioners may assume that service users are ‘asexual’ [65,42,24], and relegate issues of sexuality and romance to a position of low priority for this reason.

### Methods of relationship support: current practice and suggestions

Staff respondents discussed a range of methods of support, including preparing the service user by educating them on dating safety, building relevant social skills, and direct methods of assistance such as dating app coaching or assisted socialising. In other social domains such as finding employment and housing, direct methods seem to have more support in the literature, and working firstly on a service user’s ‘readiness’ to engage is found to be unhelpful [66–69]. Moreover, regarding dating sites, it has been reported that using such online means makes it easier for those with mental illness to screen out inappropriate partners and locate a smaller pool of potential partners from a safe distance [32,70].

Furthermore, one documented skills group had accompanied social outings written into its manual, where service users were able to learn and develop social skills in a real dating environment, and afterwards discuss their experiences with their group leader, which service users found both acceptable and effective [54]. This literature tentatively suggests that direct methods of support may be preferable. However, due to a lack of literature comparing the effectiveness and acceptability of indirect compared to direct methods of support, this conclusion cannot be made without additional research.

The staff respondents also mentioned ‘discussions’ factoring in their current practice, and as a suggestion to improve future practice. In the literature, there exists some guidance surrounding the communication of sexual issues, which could potentially be extended to relationship seeking conversations also. For instance, both the PLISSIT [71,72] and BETTER [73] models advocate addressing topics of sexual wellness for disabled and ill patients in medical environments. These models have in common the following structure: (1) raising the topic of sexuality (2) explaining that sexuality is part of the quality of life (3) telling the service user about resources available, and (4) conveying their capacity in addressing concerns and questions [74]. Such a guided approach may have potential to help reduce staff hesitation about broaching this conversation.

### Strengths and Limitations

A strength of this study is that all responses were provided anonymously and remotely. This means there was minimal social desirability bias, and participants could feel able to present any and all personal opinions without fear of judgement. Using an online survey as opposed to interviews meant there was more scope to hear from a wider range of staff working in a variety of roles in different mental health and social care services.

However, some limitations must also be acknowledged. Firstly, typed free text responses elicited fairly brief responses from respondents. It was not possible to probe for more information, as would be possible within a semi-structured interview. This resulted in some responses being difficult to interpret.

Secondly, the survey achieved a relatively small number of responses, precluding any correlation analyses and/or inferential statistics to investigate associations between clinicians’ opinions, and the job roles and type of service that they worked in. Larger studies could inform whether certain roles or services need specific training, or more training than other roles or settings. The small sample size also means that the current results must be interpreted with some caution; those who were more interested in the topic may have been more likely to respond. This is also a limitation of convenience sampling methods such as snowballing. In addition, only 15% of respondents were male. Although this reflects the national gender proportions of staff working in mental health [75], there was some suggestion from our results that male staff may have felt less comfortable in having ‘finding a relationship’ conversations in general, and especially with female service users. Further qualitative and quantitative studies are indicated, engaging with a broader range of stakeholders, including service users and service commissioners.

### Implications for practice

The first implication of the current study is the recognition of a need for acknowledgement and explicit support from mental health services and policy makers as to the importance of this topic. Clear and explicit guidance to staff about whether offering relationship seeking support is endorsed in the service is desirable. Services could also helpfully provide guidelines within service policy, alerting staff members to both what is expected of them regarding ‘finding a relationship’ conversations, as well as standard rules of conduct [24].

In addition to supportive policy, guidance or training for staff on the topic of supporting service users in regard to romance and intimacy is desirable. Training could able to address multiple issues raised by the current participants, such as discomfort, perception of inappropriateness and feelings of being ill-equipped. As suggested by the current participants, such training could increase knowledge about relationship and intimacy needs in mental illness, enhance skills to talk to service users about these topics comfortably, and most importantly, perpetuate the attitude that this is an integral aspect of psychological practice [49]. They would also address complex areas such as service user capacity and consent. Such training programmes may be informed by existing campaigns in the field of learning disabilities, such as *Supported Loving*, which educates both professionals and stakeholders on the importance of healthy romantic and sexual relationships for service users. The widespread success of this campaign underscores the equivalent need for such resources in the field of mental health [76].

## Conclusion

In conclusion, there appears to be willingness amongst mental healthcare staff to increase the provision of relationship seeking support to service users. While there is much hesitation, some of this may stem from the unfamiliarity of this topic area in mental health and social care services. Therefore, it is recommended that staff guidance, training programmes and ways of working are developed and evaluated. These should be informed by future in-depth qualitative research with service users and staff, to educate and improve staff confidence and skills on appropriate methods to support service users who wish to seek an intimate relationship. Such resources may empower mental healthcare staff to have open, non-biased discussions with service users, and to implement direct methods of relationship seeking support, such as supporting with accessing dating sites, social opportunities, and dating skills groups. Overall, this study has identified a need for more research and work in this area to encourage staff to have conversations with service users about finding intimacy, in the interest of achieving important, but currently neglected goals for service user recovery and quality of life.

## Declarations

### Ethics approval and consent to participate

This study was assessed and approved by UCL’s Research Ethics Committee (Project ID: 24833/001).

### Consent for publication

Consent was obtained at the commencement of each person’s participation in the online survey.

### Availability of data and materials

All data generated or analysed during this study are included in this published article [and its supplementary information files].

### Authors’ contributions

AER completed the literature review, and the collection and analysis of data. BLE, HK and SE all contributed to the editing of various drafts.

## Data Availability

All data produced in the present work are contained in the manuscript

## Acknowledgements

Thank you to my three supervisors, Brynmor Lloyd-Evans, Helen Killaspy, and Sharon Eager, without whom this project would not have been possible.

## Appendix A Online survey including participant information documents

Researcher contact details:

MSc Researcher: Angelica Emery-Rhowbotham; Principal investigator: Prof. Brynmor Lloyd-Evans.

Thank you for helping with this project which is being conducted by an MSc student in the Division of Psychiatry at UCL. Previous research at UCL has found that mental health service users who do not have an intimate / romantic relationship would often like to find a relationship but that staff and service users perceive some challenges with knowing how and when to talk about this and what help could be offered. The aim of this study is to learn from mental health and social care staff about how and when they talk to service users about finding a relationship, what barriers arise in discussing this, and suggestions for ways to help.

### Who are we looking for?

We are seeking mental health and/or social care staff (whether working for the NHS, Local Authorities or voluntary sector services) to complete a brief online survey to tell us your views and experiences on this topic.

If you work in a specialist sexual health, relationship counselling, psychosexual therapy or Gender Identity service, please do not complete this survey. We are keen to understand the average occurrence of “finding a relationship” conversations in mental health settings. As conversations about relationships are well represented in the above settings, this study wishes to focus on settings where such conversations are less common.

### What happens if you agree to take part?

If you decide to complete the survey, you will be asked a number of questions on your experience and opinions about helping service users to find a relationship. We will also ask you for some information about you: your age, gender, ethnicity and job role. The questionnaire will then ask you about: how you currently engage in helping service users to find a relationship; barriers and facilitators you experience in having “finding a relationship” conversations; training and other support you have been given in this area, as well as your opinions on particular methods of helping service users to find relationships.

These data will be kept anonymous with no way of tracing them to you. You can complete the survey anonymously, without leaving your name. This means that we as a research team will not know who has completed the questionnaire and who has not. However, if you wish, you can leave us your email address so the research team can send you a report of the results from the completed study, and, if you wish, contact you about taking part in a more in-depth interview in the future. If you decide to provide your email address for follow up and future contact, this will be stored securely and separately to your answers so that your data will remain anonymous.

The survey will take up to about 15 minutes to complete. You can quit the survey and leave it unfinished at any point if you wish and your responses will be deleted. However, please note if you would like to revisit the survey again, you will have to restart the questionnaire from the beginning. Because the data are anonymised, it is not possible to withdraw your data once you have completed the survey.

We hope this work may result in increased understanding of ways to help service users to find a relationship. We hope it may also increase understanding about what support mental health and social care staff need in having “finding a relationship” conversations and addressing service users’ needs for relationships.

Findings from this survey will be written up by the lead researcher for her MSc dissertation in September 2023, and will then be submitted for publication in a scientific journal. A report summarising findings from the study will be sent to all survey respondents who choose to leave a contact email.

This research project has been reviewed and approved by UCL REC. Ethics ID number: 24833/001.

### Local Data Protection Privacy Notice

The controller for this project will be University College London (UCL). The UCL Data Protection Officer provides oversight of UCL activities involving the processing of personal data, and can be contacted at

This ‘local’ privacy notice sets out the information that applies to this particular study. Further information on how UCL uses participant information can be found in our ‘general’ privacy notice: please click here.

The information that is required to be provided to participants under data protection legislation (GDPR and DPA 2018) is provided across both the ‘local’ and ‘general’ privacy notices.

The categories of personal data used will be as follows:

Age

Gender

Ethnicity

Religion

Job title

How long you have worked in mental health or social care

Professional group

Type of service you work in

Type of sector you work in

The lawful basis that would be used to process your personal data will be performance of a task in the public interest.

The lawful basis used to process special category personal data will be for scientific and historical research or statistical purposes.

Your personal data will be processed so long as it is required for the research project. We will only be able to link these personal data to you if you choose to leave us an email address, otherwise the survey is complete anonymous at source. We will anonymise all personal data you provide us with, and will endeavour to minimise the processing of personal data wherever possible. Your email address (if you have chosen to provide it) will be kept securely for 12 months after your participation, at which point it will be destroyed. Other personal data will be archived for 10 years for the potential use of other researchers, at which point that too will be destroyed.

If you are concerned about how your personal data is being processed, or if you would like to contact us about your rights, please contact UCL in the first instance at.

### Contact for further information

If you have any questions or concerns, please contact Angelica Emery-Rhowbotham (MSc researcher) at or Prof. Brynmor Lloyd-Evans (Principal Investigator) at.

If you feel that your concerns have not been adequately addressed or resolved by the research team, please escalate your concerns to UCL REC at.

Please tick here to confirm you have read this information and consent to take part in this survey on this basis:

### Online survey

Thank you for taking part in this study. What follows are some questions on the topic of relationships, specifically staff perspectives on “finding a relationship” conversations between mental health and social care workers and service users.

In “finding a relationship” conversations, a service user and staff member discuss the prospect of the service user finding an intimate / romantic relationship. A relationship may include a singular partner, or other type of dynamic such as a non-monogamous or polyamorous relationship. We are interested to discover the context and content of such conversations, as well as any barriers perceived by staff in having these conversations. The following questions should take up to 15 minutes to complete.

Please take care not to reveal any identifying details or personal information about service users you may be thinking of. This is in respect of their privacy.

After you have answered each question, press the ’next’ button at the bottom of the screen to move to the next page. Please note once you have progressed to the next page, you will not be able to revisit or edit your previous answers.

Thank you for taking part in this survey.

Click the arrow to start the survey now. ->

#### Part 1 – About you

1. How old are you? - 18 - 25 - 26 – 35 - 36 – 45 - 46 – 55 - 56 – 65 - Over 65 - Prefer not to say
2. Which of the following best describes your gender? - Male - Female - Other (please specify) - Prefer not to say
3. Which of the following best describes your ethnicity? Asian, Asian British / Black, Black British, Caribbean or African / Mixed or multiple ethnic groups / White / Other (please specify) / Prefer not to say
4. Do you follow a specific religion? Buddhism / Christianity / Hinduism / Judaism / Islam / Sikhism / Other (please specify) / No religion / Prefer not to say
5. How long have you worked in mental health services? Less than 2 years / 2-5 years / 6-10 years / More than 10 years / Prefer not to say
6. What is your current job title? (If you would rather not say, please leave this blank). [Free text response]
7. Which professional group, if any, do you belong to? Social worker / Occupational therapist / Psychologist / Psychiatrist / Counsellor, therapist / Support worker / Peer support worker / Other (please specify) / Prefer not to say
8. Which of the following, if any, do you work for? NHS / Local authority / Voluntary sector organisation / Independent sector organisation / Prefer not to say
9. What sort of service do you mainly work in? NHS community based mental health team / Day service (e.g. day centre, drop-in service, recovery college) / Supported accommodation service (e.g. residential service, supported housing, floating outreach) / Inpatient service / Other (please specify) / Prefer not to say
10. Do you work in the U.K. or elsewhere? - U.K. / Elsewhere (please specify)

#### Part 2. Talking to Service Users

11. Some service users express that they would like help from services in finding and developing intimate / romantic relationships. How far do you agree that this is an appropriate aspect of your work role? Strongly agree / Somewhat agree / Somewhat disagree / Strongly disagree
12. Please explain briefly why you think this is or is not part of your work role. [Free text response]
13. Of the service users you support, how many are single? (Your best estimate is fine.) Less than 25% / 25-50% / 50-75% / More than 75%
14. How many of these single service users would like to find a relationship? (Your best estimate is fine.) Less than 25% / 25-50% / 50-75% / More than 75%
15. Of the service users you support who are single, with how many do you have conversations about “finding a relationship”? None / Few / Many / All or nearly all
16. How often do you have “finding a relationship” conversations with the service users you support? Never / Rarely / Sometimes / Frequently
17. Who initiates these conversations? - Always the service user - Usually the service user - Sometimes me sometimes them - Usually me - Always me - Not applicable – I don’t have these conversations

#### Part 3. Exploring barriers

18. How much do the following factors form barriers to “finding a relationship” conversations?

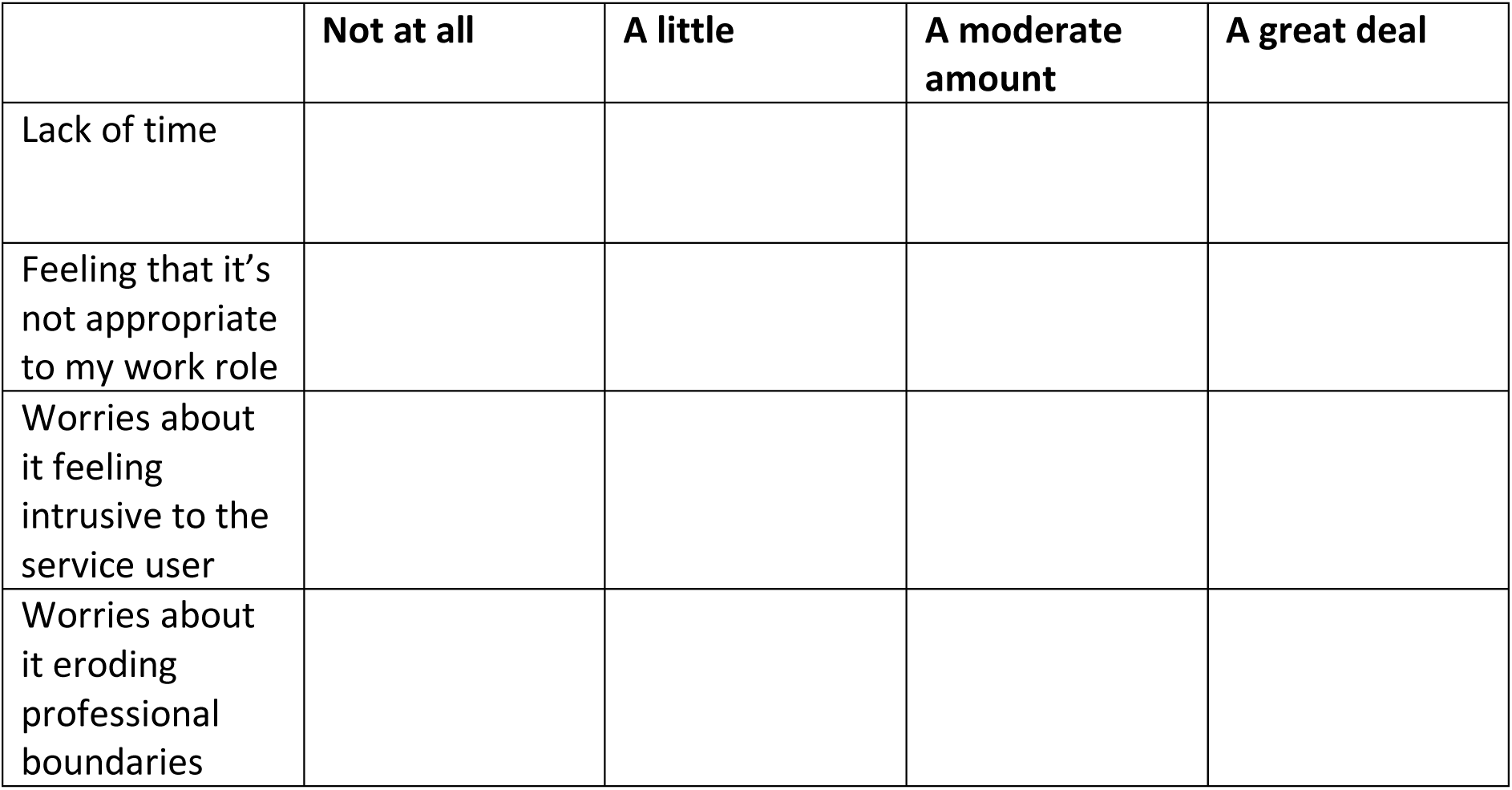
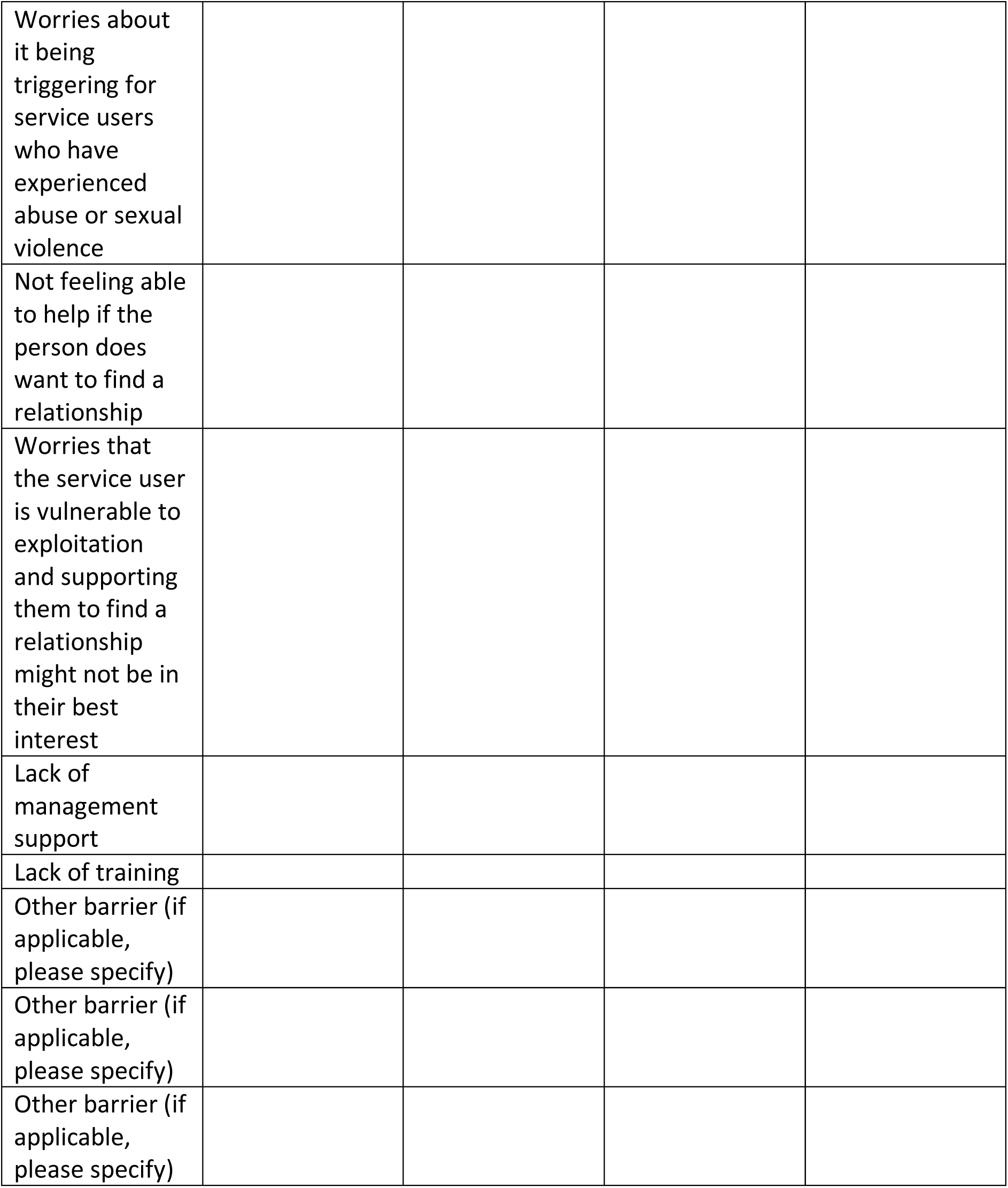
19. Which of these barriers do you think is the most significant? [Free text response]
20. Is there anything else you’d like to say about these barriers? [Free text response]
21. In which context, if any, is it easier to have “finding a relationship” conversations? - One-to-one conversations - Group settings - Other (please specify) - Unsure
22. Please tell us if any of the following service user characteristics affect how comfortable you feel having “finding a relationship” conversations with service users? (please tick)

23. If you selected any of the factors above, please briefly explain how these affect how comfortable you feel having “finding a relationship” conversations. [Free text response]
24. Please tell us about any ways in which you, or others you work with, try to help those who express a desire for an intimate / romantic relationship. [Free text response]
25. Are there any other ways you think staff in mental health and social care services could support people in finding a relationship (even if these are not current practice in your workplace)? [Free text response]
26. Have you been given any guidance or training in your current work about having “finding a relationship” conversations with service users? - Yes / No 26a *If yes* Please tell us briefly about what guidance or training you have had regarding “finding a relationship” conversations. [Free text response]
27. Are you aware of any organisations which provide examples of innovative practice in supporting service users who wish to find a relationship? If yes, please briefly tell us about them. [Free text response]
28. How <u>appropriate</u> do you think it would be to provide the following types of support in your service? [Tick box matrix]

|  |
| --- |
| **Gender** |
| **Religion** |
| **Age** |
| **Ethnicity** |
| **Sexuality** |
| **Level of social skills** |
| **Other (please specify)** |
| **Unsure** |

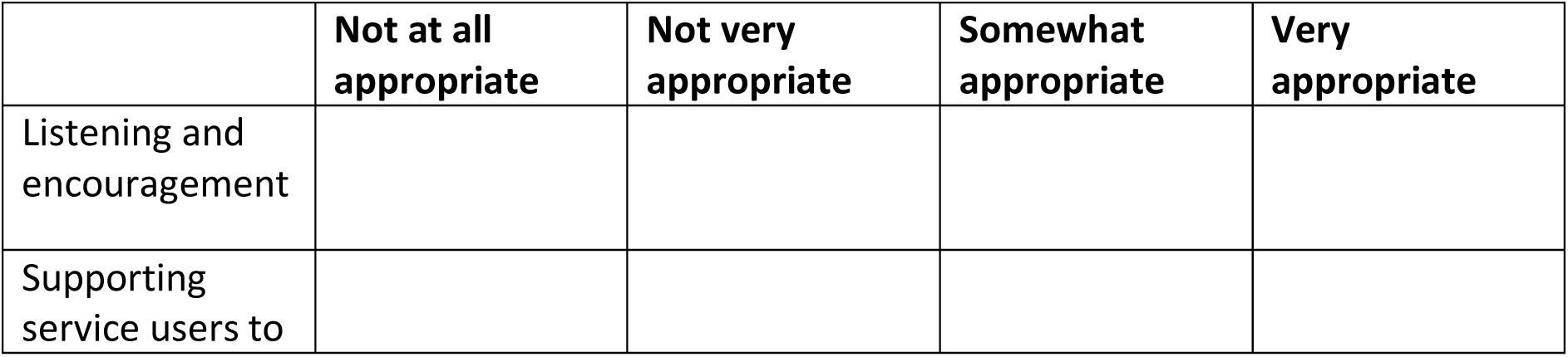
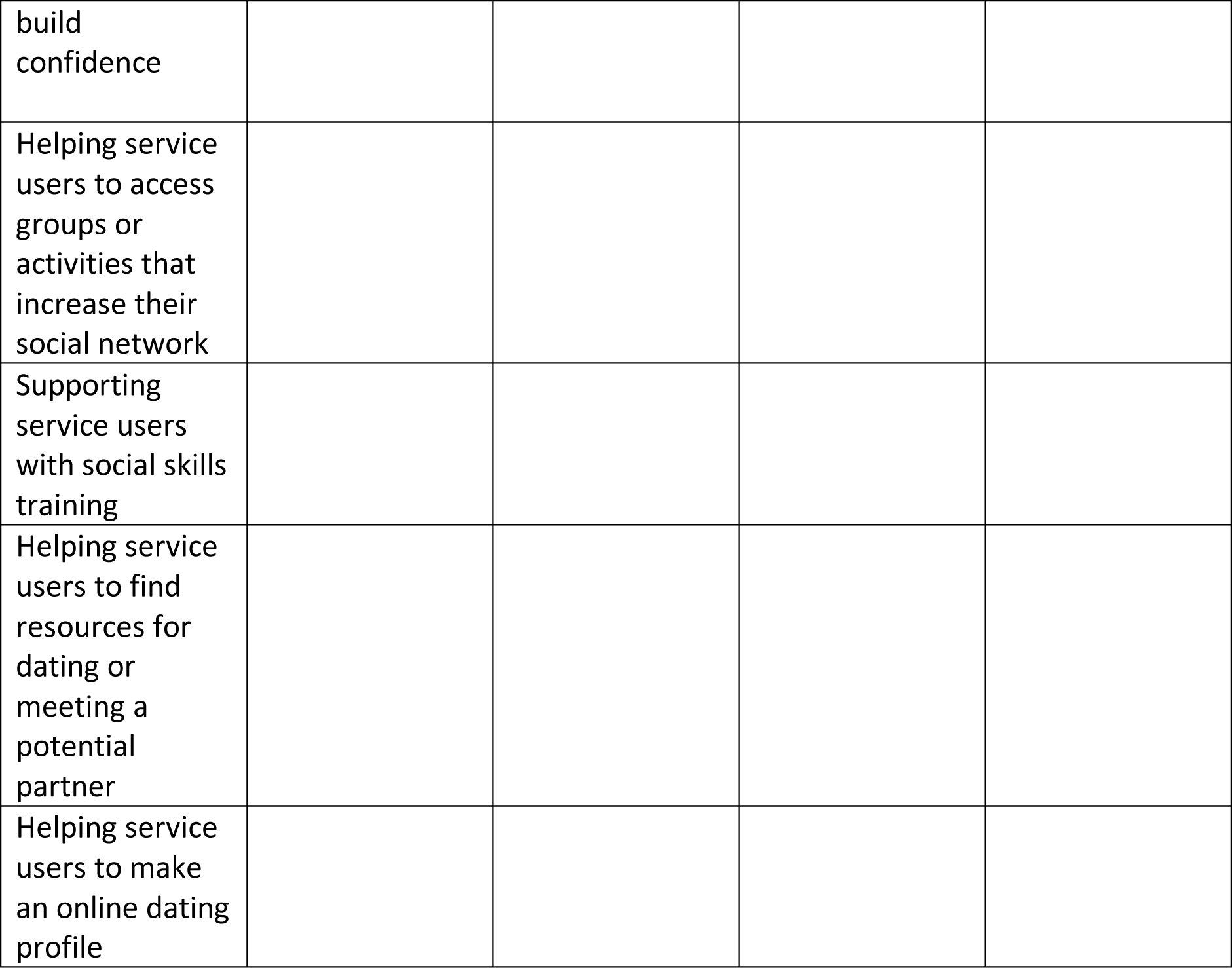
29. How <u>feasible</u> do you think it would be to provide the following types of support in your service at the moment?

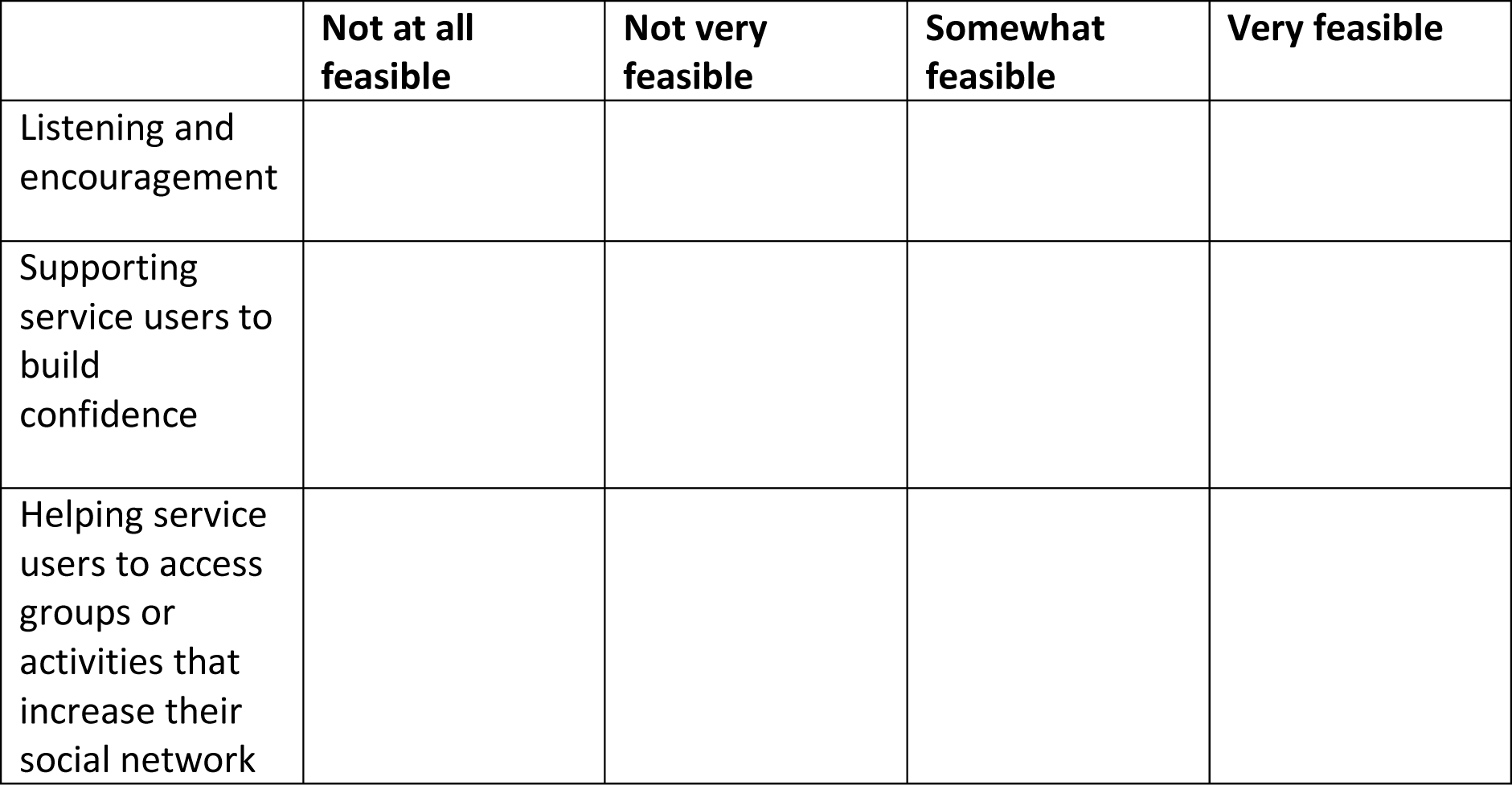
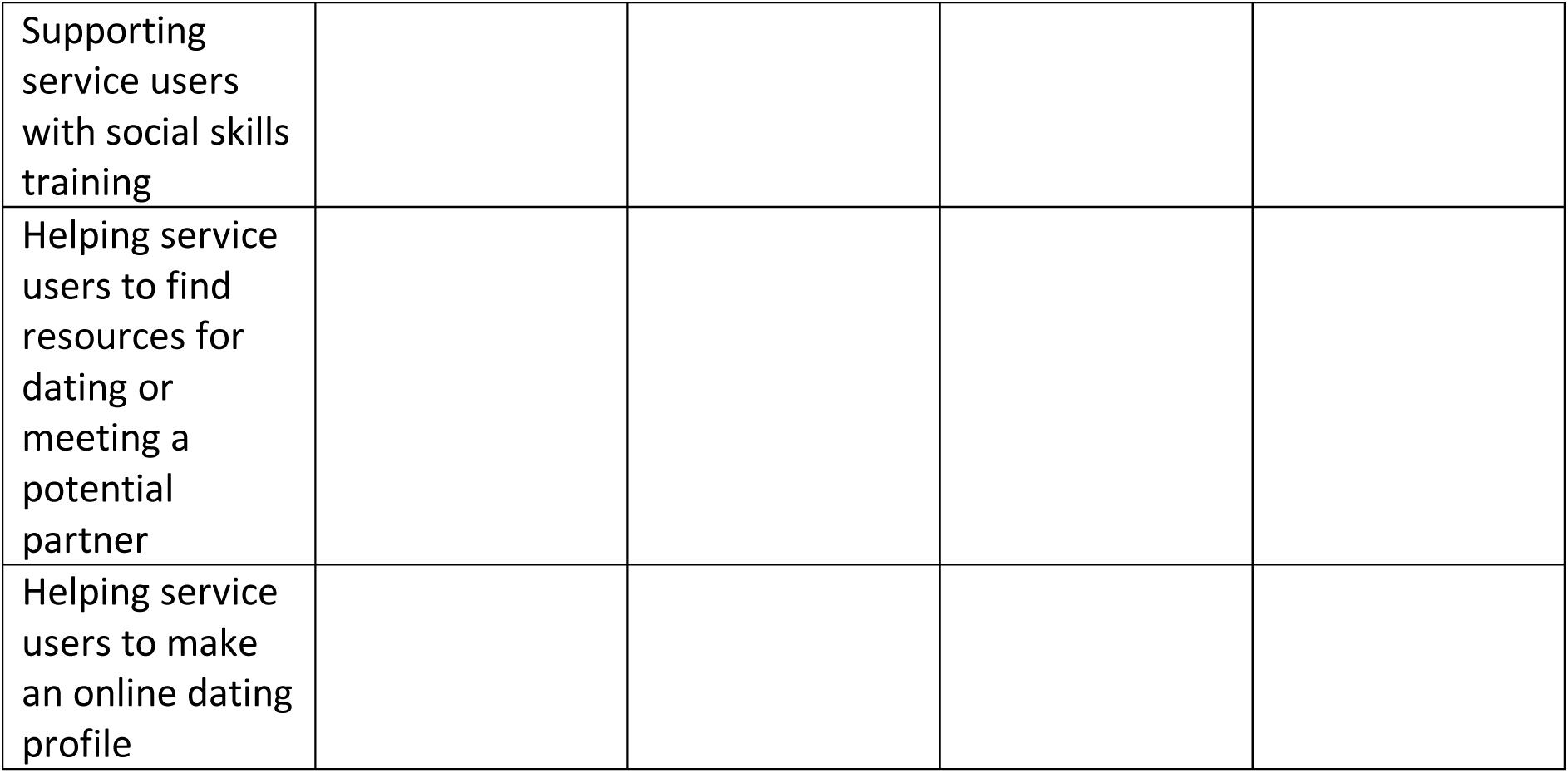 Thank you for taking part in this study.
30. Would you like a report of the findings from this survey? - Yes / no
31. May we keep your contact details to ask you if you would be interested in participating in any future research by our research group on this topic (e.g. an in-depth interview)? - Yes / no
32. Could we contact you to ask about sending us a copy of your organisation’s policy on talking to service users about finding a relationship? - Yes / no If you answered yes to any of the above questions, please provide us with your email <u>by</u> <u>clicking on this link.</u>

## Appendix B Coding process for free-text responses

### Step 1: Creating initial themes for each of seven qualitative questions

#### Question 1, Appropriateness: ‘Please explain briefly why you think this is or is not part of your work role.’

##### AGREE

Part of holistic care or recovery

- Part of holistic care planning [6]

- Part of recovery and happiness [10]

- My patients talk a lot as part of their recovery is ‘finding love’ [23]

- It’s a key part of people’s lives and recoveries. But does require some delicacy to do well and some evidence on how to do it well would be good too. [26]

- Relationships are an important aspect of overall wellbeing, and health. Whilst we may not be experts at supporting people in forming intimate relationships, or indeed matchmaking, our role should include an interest in all aspects of a person’s emotional health - and they should be made to feel comfortable discussing such matters. Just as their physical wellbeing, or wishes regarding employment/education etc are all potentially relevant to their mental state. [30]

- its important for the individual’s wellbeing, QoL etc. It can however be difficult to support, and in some instances need to considering safeguarding risks. [27]

- OTs work holistically with all aspects of life [36]

Identity or role fulfilment

- It is a vital part of identity and role fulfilment [14]

Service user’s desire

- Having a meaningful relationship with someone is many peoples goal. it is very much my role to help/support/advise in ways I can [15]

- this is a pressing concern and need in the service users I support [24]

Good for mental health

- Romantic relationships are key to managing mental health and to manage lonliness [16]

- I feel that romantic relationships and intimacy are a human need and have a huge impact on mental health. [25] Recognising healthy relationships and how to behave in them

- but we should support our service users to recognise healthy relationships, build boundaries and encourage positive social interactions [9]

- To support them to think about what a healthy relationship is and to think about the way in which they meet new people. [19]

Self improvement support (indirect support)

- I would support with self-improvement goals to help the chances of finding a relationship e.g. healthy living, physical fitness, employment, hobbies [13]

Skills building helps therapeutic alliance

- however, helping them to develop appropriate skills would help to build therapeutic relationship and may result in better mental health outcomes for them. [3]

Only if MH related

- If they are struggling to maintain or find a relationship based on MH difficulties then i understand that our role can help them with self-esteem or any social anxiety but i think its only our work role if its MH related, not just because someone without MH cant find a relationship. Not sure how we are suppose to help there [29]

- I would see supporting someone to develop their skills and knowledge around navigating romantic relationships as a part of my role if important to the person. Particularly if mental health difficulties or learning difficulties or neurodivergence was a barrier to this.. [34]

Relevant therapeutic goal

- Patients are concerned regarding body image and struggle with intimacy [17]

- I think it should be more because it is a strong motivator and some patients do not have experience of developing relationships of any kind [32]

- Depression can be linked with loneliness and a lack of connection with others.

Helping someone to try dating again could be really beneficial to their mental health. [33]

Service users want support

- I think people want this but do not express it as they dont feel it is within our role [18]

Considering needs and roles within relationship

- As a psychologist, I believe we have some capacity to help people to consider their needs and roles in relationships. [21]

Needs boundaries

- I think as long as you keep it within certain boundaries, why shouldn’t you support them? [22]

- Whatever the occupational barriers might be, i would explore goals and ways of overcoming them. There would be limits however. [37]

Help to access potential partners

- We can help people attend groups etc or meet new people but we can, Äôt really promote anything that isn, Äôt regulated. Would be a challenge to help them find this relationship [38]

##### DISAGREE

Unethical

- I feel like it would be unethical to help them find a relationship, [3]

Work role is closely delineated

- My work role involves logging into an app at the start of the shift, and ticking off the tasks that are listed e.g. prepare food, so the work is mostly set out for us. We do help assist with emotional support and distress, but no tasks are really geared towards building relationships. [5]

-). I also do outreach as part of my job with an autistic male who is more high functioning, and has said once that he seeks a relationship. My role when I started was to encourage him to try new activities in London and discuss any emotional difficulties if they arise (such as struggling with depression), and I think that is the role I feel responsible for as opposed to helping them find a partner. [5]

Not something clients desire

- Additionally, the clients have severe mental/physical disabilities and so it is either not something they would desire (e.g. not mentally capable of), or [5]

Service users not capable of social interaction

- they are too severe to interact with the public/other society members e.g. prone to anger and attacking (e.g. not socially capable of) [5]

Not something service users bring up

- I think that is also how they view me so he wouldn, Äôt necessarily go to me for that (possibly because they don, Äôt deem as appropriate) unless I approached the topic. [5]

Abusive clients

- Many of the clients have history of being perpetrators of DV. [7]

- There are also risks to be considered, for example of people ending up in abusive relationships/one with very unequal power dynamics. [26]

Lack of time and resources

- Even though this is important, it is difficult to address these issues in the time slot given to me to see the service user. I also do not have access to resources or skills to be able to help someone like that. [8]

- Also, there is barely enough time to do the core aspects of my job, so in my opinion there is very unlikely to be resource for this. [31]

Can’t be matchmakers

- We cannot act as matchmakers [9]

Inappropriate

- I’m not sure how appropriate helping someone form an intimate relationship is? [12]

- It feels slightly intrusive to become involved in someone’s romantic life; a breach of a professional boundary. [31]

Professional responsibility

- I would support with self-improvement goals to help the chances of finding a relationship e.g. healthy living, physical fitness, employment, hobbies, but not with actually finding a relationship as there is a concern about professional responsibility if something goes wrong

Safeguarding risks

- It can however be difficult to support, and in some instances need to considering safeguarding risks. [27]

Relying on other professionals

- tend to rely on support workers to help them socialise but can suggest this as part of the care plan [28]

- However, perhaps social workers / OTs within my team could run groups on safety / appropriate when online dating etc. which would be in keeping with this aim. [31]

Relationships are relevant, but not actually finding a partner

- It, Äôs relevant to mental health, and somewhat relevant in terms of values and behaviour, but actually practically finding somebody feels a little beyond our scope [35]

##### IT DEPENDS

Depends on their level of care: high support not priority

- It depends on the level of care they require, in the more independent housing I would say I would be happy to discuss this but in high support this is not our priority [4]

Undecided

- I’m undecided I think it could be good for residents to find meaningful relationships but I wonder if it crosses a boundary to support them into those relationships [20]

#### Question 2, Barriers: ‘How much do the following factors form barriers to “finding a relationship” conversations? - Other barrier (if applicable, please specify) – Text’

Relationship being detrimental to the service user

- it is probable that a relationship may present another stressor to the service user’s life (moderate amount) [4]

Service user not capable of relationship

- A view that the individual may not be socially capable of a relationship e.g. if they display traits of aggression (mod; 7)

Service user may put their partner at risk

- The clients who have asked are male and have a history of perpetrators of domestic violence [9]

- Might be a DV perpetrator themselves so this comes with risks [31]

Service user not asking for support

- The person who we support being able to properly communicate their desire for a relationship even if it is something they might want [17]

Service user not ready for a relationship

- just had a baby, might not be ready yet. [36]

Liability

- Worry that if it goes wrong will the resident blame me? [37]

Getting it wrong

- If a person has a severe learning disability and has always been single, conversations about relationships (if handled incorrectly)may come across as condescending or as if you are making fun of them [17]

Being ill equipped

- Also feel inequipped like it would be opening a tin of worms [50]

Other people are better suited to this work

- I think I’m not necessarily the best person as a consultant psychiatrist carrying out relatively formal reviews - conversations better initiated by people like care coordinators or support workers in a relatively informal setting. [44]

Not a priority issue

- Other management priorities [50]

- Needing to prioritise more pressing current issues (e.g. risk, accommodation, getting out more in geberal). Sometimes other things need to be in place first. [51]

Related barriers

- Other support around the person such as family or services being opposed [52], a little

- service users’ previous relationship problems [4]

Colleague hesitation

- Colleagues opinions of whether it’s our role [55]

Service organised differently

- Relationship goals not being part of routine assessment [52]

Not appropriate context

- starting up relationship problems in a therapeutic session [4]

Families being overprotective

- If the family are active in the person we supports lives, they could be overprotective and not want their child to have a relationship e.g. they feel they are too vulnerable or would not want staff to be involved in that way [17]

- If the family are heavily involved, they can often be overprotective and not want their child to be dating [7]

Needing therapeutic alliance first

- People not wanting to discuss this until they have built a trusting relationship with the professional [52], mod

Stigmatising singlehood

- also concerns that the discussion (where initiated by clinician) stigmatise their single status) [46]

#### Question 3, Barriers: ‘Which of these barriers do you think is the most significant?’

Inappropriate in my work role

- feeling it is inappropriate in my work role [P2]

- Worries that is inappropriate and intrusive [P4]

- Concerns re: relevance/appropriateness - not personally, but at service level and service user level. [P51]

Breaking boundaries

- Worries about it breaking professional boundaries [P9]

- Ensuring the support remains boundaries and the service user takes the lead if this is what they wish to do [P10]

- professional boundaries [P17]

- Professional boundaries, [P19]

- The barrier around professional boundaries [P31]

- crossing professional lines [P35]

- Boundaries [P51]

- Boundaries, triggering and intrusiveness [P58]

Intrusive

- Worries that is inappropriate and intrusive [P4]

- worries it might seem too intrusive [P41]

- Being intrusive to the service user. Also, a make worker asking a female might be viewed dimly or inappropriate. [P49]

- Boundaries, triggering and intrusiveness [P58]

Gender issues between client and clinician

- Being intrusive to the service user. Also, a make worker asking a female might be viewed dimly or inappropriate. [P49]

- Nature of the service and being a male member of staff I would not feel comfortable discussing with a female patient and would also be concerned that this would make them uncomfortable. [P54]

- I think gender of staff member [P54]

Relationships not prioritised

- Prioritising other issues [P45]

Lack of training

- Lack of training [P32]

- Lack of management training [P13]

- I have experienced no training or discussions around this therefore naturally you think it might be out of the scope of your professional boundaries. Something about it does feel that way. [P25]

- Lack of training around conversations and how these can be appropriate [P36]

- Lack of training perhaps [P42]

- Lack of training [P47]

Hard to identify clients

- Hard to identify just one and varies from person to person [P38]

Lack of time and resources

- Time [P8]

- Time and appropriate services [P12]

- Time, and feeling unable to help. [P43]

- Time constraints are a factor [P27]

Low clinician confidence

- voulnerability. I’m and experienced nurse although low confidence with this [30]

- not knowing how to support someone to seek a relationship and worry about their vulnerability [P24]

- Time, and feeling unable to help. [P43]

- Not feeling able to help [P57]

- Not being able to do anything to help if they want a relationship [P59]

Client’s history

- Client’s history of domestic violence [P7]

- Sexual assault [P41]

Client context

- Either there pregnant or have a baby up to the age of 2. May need support for this before getting into a new relationship. [P30]

Vulnerable patients

- vulnerable patients [P19]

- voulnerability. I’m and experienced nurse although low confidence with this [P24]

- not knowing how to support someone to seek a relationship and worry about their vulnerability [P27]

- Patient being vulnerable [P34]

- Sexual assault [P41]

- worries they will be exploited, or exploit someone [P57]

- Boundaries, triggering and intrusiveness [P58]

- worries about service user being vulnerable to exploitation [P62]

Support

- Lack of management support [P5]

- Management support and the general perception of the public about people with mental illness dating [P33]

- Other supports and services seeing this as unimportant or inappropriate [P46]

- Management support [P48]

- Lack of support / training around having these conversations [P56]

- nature of service are largest factors [P54]

- 

Training

- I have experienced no training or discussions around this therefore naturally you think it might be out of the scope of your professional boundaries. Something about it does feel that way. [31]

- Lack of support / training around having these conversations [P56]

Societal perception

- Management support and the general perception of the public about people with mental illness dating [P25]

- I would not want to ask someone about finding a relationship if they have not brought it up themselves. This is because I would not want to assume that finding a romantic relationship is something they need or want to do - I don’t want to impose societal expectations of relationships on someone, particularly if struggling with their wellbeing. It would need to be worded more broadly, i.e. are you interested in relationships / is this something that’s important to you? [P51]

None

- None, it is part of the role to address this aspect of their lives as well in thinking about the person as a whole [P6]

#### Question 4, Barriers: ‘Is there anything else you’d like to say about these barriers?’

Management and policy

- I think the main barrier is management, it is never really spoken about that we need to help the clients find relationships, especially because there are so many people involved that it would need to be discussed with e.g. the care management, social workers, the family. When we’ve had less severe clients in the past who weee definitely capable of relationships and I think would benefit/enjoy it, it was something that was never spoken about or we were trained in /encouraged to help so it was never something we tried to talk about with them. [17]

- I understand their importance but residents do want someone in their life romantically and I want residents to feel happy it would be helpful if there were more clear guidelines on how we can support a client with this [37]

- Everything you do has to be mostly approved or encouraged by them, so even if you have no personal barriers, without management support, it is not something that can be done. [7]

Staff need training

- more training in this regard [41]

- they are very daughting so would i feel staff would like support arund this [33]

Staff may get it wrong

- It could appear as though single staff members are flirting if this conversation was not done effectively [56]

Important issue

- it is very important that this topic is spoken about, this is huge in peoples lives [30]

- It seems a shame because romantic relationships are so important and could be the changing factor in someone’s loneliness, suicidality and wellbeing. [31]

- This is a very important area, and I welcome a better understanding of how to navigate these issues and offer a more holistic approach to clients [48]

Client needs to be stable before engaging in relationship

- Need to focus on own mental health and wellbeing and care of a baby. Get to know other people but take things slowly if it works out they will understand. having a baby is one of the most difficult things you’ll ever do as a women, and if someone is unwell some awful individuals will take advantage of this. To think about where, when and why your meeting someone new, making friends and family aware so that they know if you do get into any bother. [36]

- Sometimes people, at least in more acute services, might benefit from more stability (e.g. of mpod, of routine, being able to go out) if they are then to find a hood relationship. But this is cery influenced by the fact i wiork in secondary and inpatient services. [51]

#### Question 5, Current practice: ‘Please tell us about any ways in which you, or others you work with, try to help those who express a desire for an intimate / romantic relationship.’

Just having a conversation

- asking a simple question like “What are your thoughts about romantic relationships?” [4]

- Just having a conversation about it and finding out their thoughts. [39]

- conversation, reassurance, practical advice [41]

- Explore whether they have any pre-existing ideas in mind first and take it from there [8]

- I ask about satisfaction with intimate relationships as part of an initial screener/putcome measure. [55]

- 

Discerning client goals

- to ask whether having a relationship is one of their goals [46]

- asking what a person would like in the future or work towards [30]

Exploring relationship desires

- I have chats with my clients about what kind of relationship they want. I try to support them by asking how they like to reach out to significant others i.e. in person/online and discuss safety when dating e.g. keeping to public spaces not sharing personal information etc. more of an advisory role as opposed to engaging them directly. [37]

Discussing barriers

- I guess figure out whats stopping them in the first place or what barriers they have and figure out what we can do to help with those barriers [47]

Discussing social network

- Discussion of social connections [6]

- Optimising mental health can sometimes facilitate this, as can helping people to engage in social activities. But this is an indirect effect of something that is done anyway. [49]

Not done in my service

- This is not something that is done in my practice. [7]

- In 20 years I’ve not witnessed this [40]

Take service user’s lead

- If sex or romantic relationships are specifically raised as an identified issue, this will be explored, formulated and any barriers discussed. Intimate/family relationships is a part of the care plan, however it rarely is a priority to focus on within the time frame. [31]

- Begin to discuss when they bring it up - based around their circumstances there is no guidance [50]

Relationship education

- We try and sensitively explain how consensual relationships work. Remind them how coercive behaviour is wrong and try and guide them on acceptable behaviour [9]

- explore healthy and unhealthy relationships. To think about the whole picture in keeping themselves safe keep talking about it. [36]

- I have chats with my clients about what kind of relationship they want. I try to support them by asking how they like to reach out to significant others i.e. in person/online and discuss safety when dating e.g. keeping to public spaces not sharing personal information etc. more of an advisory role as opposed to engaging them directly. [37]

- We can provide support and encouragement, help people to identify their needs and goals, teach people about communication and relationship skills, and connect people with resources in the community. [45]

- Talking about and having easyread materials available describing qualities of supportive romantic relationships, and dating safety including online. Role playing starting conversations or using drama or discussion to explore feelings around romance and sexuality. Also sadly safeguarding as sometimes this is expressed in the context of abusive relationships. [52]

- Meeting sexual needs in hospital, social skills, online safety [54]

Increasing access to partners

- Direct them to dating sites o encourage their families/carers to support [10]

- look into options of where to find a good match - common interests [27]

- behavioural experiments using dating apps and broadening social opportunities. [31]

- We can provide support and encouragement, help people to identify their needs and goals, teach people about communication and relationship skills, and connect people with resources in the community. [45]

Skills building

- It wouldn’t be about finding a relationship for them but more so about building interpersonal social skills and recognising how they might be vulnerable to exploitation [13]

- Meeting sexual needs in hospital, social skills, online safety [54]

Building self-esteem

- Building up self-esteem [15]

Therapeutic interventions

- Motivational interviewing around meeting others [31],

- Through therapy, clarifying what they want, problrm solving how they might go about it, challenging anxieties or negative thoughts about how it might work out. S [51]

- In LD service we specifically had a sex and relationships group. It comes up in CAMHS but feels less focused on as a part of the work [53]

Showing support for client’s initiative

- i show my support for them going out with friends/to pubs and using dating apps but havn’t helped practically [33]

- I have chats with my clients about what kind of relationship they want. I try to support them by asking how they like to reach out to significant others i.e. in person/online and discuss safety when dating e.g. keeping to public spaces not sharing personal information etc. more of an advisory role as opposed to engaging them directly. [37]

- conversation, reassurance, practical advice [41]

- We can provide support and encouragement, help people to identify their needs and goals, teach people about communication and relationship skills, and connect people with resources in the community. [45]

Practical advice

- conversation, reassurance, practical advice [41]

- I have chats with my clients about what kind of relationship they want. I try to support them by asking how they like to reach out to significant others i.e. in person/online and discuss safety when dating e.g. keeping to public spaces not sharing personal information etc. more of an advisory role as opposed to engaging them directly. [37]

- I would say they need to get out of their home to try and meet new people and hope they meet someone that eay [56].

Unsure

- I’m not sure! But I’m only at this service once a week so I may miss a lot of these conversations. I have spoken to clients about past relationships, but not about seeking out future ones since no desire was expressed and there were more pressing matters to address. [38]

Discussion of sexual needs

- Meeting sexual needs in hospital, social skills, online safety [54]

#### Question 6, Future suggestions: ‘Are there any other ways you think staff in mental health and social care services could support people in finding a relationship (even if these are not current practice in your workplace)?’

Attending activities outside the service

- Attending group activities in the community, more opportunities to interact with people outside of the service [6]

- Taking clients to community events e.g. coffee events with other people with disabilities, social dance classes etc. where they can be encouraged to interact with other people in society and potentially form relationships. [7]

Direct support: dating sites

- Maybe helping with dating sites but again i wouldnt know how to support someone to not get taken advantage of here. [33]

- I think it would be good to find dating services (free) that is like speed-dating or friendship making. Or provide this within the service or in the wider organisation so that residents can establish positive relationships [37]

Facilitation within the service

- Social opportunities in services [15]

- Possibly some form of safe dating facilitation - many people who use mental health services have relationships with others who do too. Quite a few fears though about the possibility that resulting relationships might turn out to be problematic in some way.

Accessing social opportunities

- Unsure, it also feels quite strange to offer a specific intervention around it as you can feel like a dating service and it might feel forced. Where as taking a social intervention and allowing any relationships to come naturally seems the more acceptable way of doing it. [31]

- I suppose via signposting to community activities? [38]

Asking about barriers to relationships at assessment

- But explicitly asking about thoughts and barriers to finding romantic relationships should be more prominent in assessment questions and interventions around mental health.. [31]

Group work

- Discussion and group work [32]

- Offer support in the form of groups to discuss what a healthy relationship looks like.

Do some scenarios and explore what they would do in that situation. [36]

- Possibly, groups to discuss online dating safety etc. [49]

Open discussion

- Discussing it more openly and not stigmatising [39]

- to give permission to patients to state this as a goal and set out how you might support someone in their socialisation to address this [46]

- Talking about sex and relationships more [54]

- Open conversion. Looking at roles and routines that might support opportunities. We can explore what the emotional, practical and esteem barriers are. There are always options but it takes the service user a lot to over come this. [55]

Psychological preparation

- And by helping to psychologically prepare them for a relationship if needed [38]

Psychoeducation

- offer some eduation about safety in relaitonships for vulnerable peope including udnerstanding their own comfortable boundaries. [33]

- Perhaps psycho Ed on healthy relationships, for some people like LD services maybe learning social skills to develop relationships [53]

Signposting

- Direct them to relevant services [10]

- Training! Being aware of organisations to signpost to that set up safe spaces for more vulnerable people to meet someone. [52]

Skills building (indirect)

- By working on the individual to build skills mentioned above that, Äôs as far as we should go [13]

- Maybe some skills support around going on dates e.g conversationa dn social interaction [33]

- Support with social skills, ensuring the person is being treated for symptoms that may affect confidence etc, or functioning, addressing sexual side effects or symptoms as well. Improving self-esteem, [48]

Needing more knowledge in this area

- more knowledge around this [27]

- I don’t know, and probably should [30]

- Training [40]

- No [41]

- Unsure [56]

Organisational change

- improving policies and access to material to meet needs [54]

Should not be part of MHP role

- I do not think it should be part of my role [9]

### Step 2: Creating overarching themes for each of seven qualitative questions

#### Question 1, Appropriateness: ‘Please explain briefly why you think this is or is not part of your work role.’

##### AGREE THEMES

###### Encouraging recovery

Part of holistic care

- Part of holistic care planning [6]

- Part of recovery and happiness [10]

- My patients talk a lot as part of their recovery is, ”finding love” [23]

- It’s a key part of people’s lives and recoveries. But does require some delicacy to do well and some evidence on how to do it well would be good too. [26]

- Relationships are an important aspect of overall wellbeing, and health. Whilst we may not be experts at supporting people in forming intimate relationships, or indeed matchmaking, our role should include an interest in all aspects of a person’s emotional health - and they should be made to feel comfortable discussing such matters. Just as their physical wellbeing, or wishes regarding employment/education etc are all potentially relevant to their mental state. [30]

- its important for the individual’s wellbeing, QoL etc. It can however be difficult to support, and in some instances need to considering safeguarding risks. [27]

- OTs work holistically with all aspects of life [36]

- It is a vital part of identity and role fulfilment [14]

- Relationships are a hugely important part of our identity and wellbeing [P51]

- We consider the social life of the person in many other ways and this is likely to be one of the most important and meaningful aspects of this for the person [P55]

- Relationships are part of well-being. It should be no different to thinking through with someone what they need for other values they have such as employment or exercise. [P61]

- our role is to help a patient holistically [P62]

Good for mental health

- Romantic relationships are key to managing mental health and to manage loneliness [16]

- I feel that romantic relationships and intimacy are a human need and have a huge impact on mental health. [P36]

- Can support mental well-being. Also important to encourage service users to think about risk/disclosure of offences as part of my responsibility to society [P57]

- We know from research that loneliness is integral to physical and mental health, and relationships are relevant to tackling isolation. [P61]

- Depression can be linked with loneliness and a lack of connection with others.

Helping someone to try dating again could be really beneficial to their mental health. [33]

###### Methods to support client relationships

Recognising healthy relationships and how to behave in them

- but we should support our service users to recognise healthy relationships, build boundaries and encourage positive social interactions [9]

- To support them to think about what a healthy relationship is and to think about the way in which they meet new people. [19]

- Discussing what a safe and healthy relationship might look like and navigating this [P59]

- 

Self improvement support

- I would support with self-improvement goals to help the chances of finding a relationship e.g. healthy living, physical fitness, employment, hobbies [13]

Skills building

- however, helping them to develop appropriate skills would help to build therapeutic relationship and may result in better mental health outcomes for them. [3]

- Agree that supporting with general social skills and areas that support development in this area is part of the role. [P59]

Help to access potential partners

- We can help people attend groups etc or meet new people but we can, Äôt really promote anything that isn, Äôt regulated. Would be a challenge to help them find this relationship [38]

###### General importance

Therapeutic relevance

- Patients are concerned regarding body image and struggle with intimacy [17]

- I think it should be more because it is a strong motivator and some patients do not have experience of developing relationships of any kind [32]

- As a psychologist, I believe we have some capacity to help people to consider their needs and roles in relationships. [21]

- I believe this is important. Because many service users can struggle to form relationships in general, and having some support regarding intimate relationship would be useful for these users. [P58]

- 

Service users want support

- I think people want this but do not express it as they don’t feel it is within our role

[18]

Service user’s desire

- Having a meaningful relationship with someone is many peoples goal. it is very much my role to help/support/advise in ways I can [15]

- this is a pressing concern and need in the service users I support [24]

###### Caveats

Needing limits

- I think as long as you keep it within certain boundaries, why shouldn’t you support them? [22]

- Whatever the occupational barriers might be, i would explore goals and ways of overcoming them. There would be limits however. [37]

- Whilst I don’t think we should be advising someone if they should/shouldnt be in a relationship. Sex and relationships are a significant part of identity and if it’s something that someone wants to explore, wants to think about, patterns, barriers etc, then it’s of course part of our job. [P56]

- 

Only appropriate if mental health related

- If they are struggling to maintain or find a relationship based on MH difficulties then i understand that our role can help them with self-esteem or any social anxiety but i think its only our work role if its MH related, not just because someone without MH cant find a relationship. Not sure how we are suppose to help there [29]

- I would see supporting someone to develop their skills and knowledge around navigating romantic relationships as a part of my role if important to the person. Particularly if mental health difficulties or learning difficulties or neurodivergence was a barrier to this.. [34]

##### DISAGREE THEMES

###### Moral and ethical issues

Unethical

- I feel like it would be unethical to help them find a relationship [3]

- psychologists and most other professionals are not trained in discussing or particularly advising around seeking/developing romantic relationships, and so it may not always be appropriate to do as it would likely be informed by subjective opinion, and could be harmful. [P51]

Inappropriate

- I’m not sure how appropriate helping someone form an intimate relationship is? [12]

- It feels slightly intrusive to become involved in someone’s romantic life; a breach of a professional boundary. [31]

Safeguarding risks

- It can however be difficult to support, and in some instances need to considering safeguarding risks. [27]

- There are also risks to be considered, for example of people ending up in abusive relationships/one with very unequal power dynamics. [26]

Professional responsibility

- I would support with self-improvement goals to help the chances of finding a relationship e.g. healthy living, physical fitness, employment, hobbies, but not with actually finding a relationship as there is a concern about professional responsibility if something goes wrong

Abusive clients

- Many of the clients have history of being perpetrators of DV. [7]

###### Not feasible in my job role

Work role is closely delineated

- My work role involves logging into an app at the start of the shift, and ticking off the tasks that are listed e.g. prepare food, so the work is mostly set out for us. We do help assist with emotional support and distress, but no tasks are really geared towards building relationships. [5]

-). I also do outreach as part of my job with an autistic male who is more high functioning, and has said once that he seeks a relationship. My role when I started was to encourage him to try new activities in London and discuss any emotional difficulties if they arise (such as struggling with depression), and I think that is the role I feel responsible for as opposed to helping them find a partner. [5]

Can’t be matchmakers

- We cannot act as matchmakers [9]

- It, Äôs relevant to mental health, and somewhat relevant in terms of values and behaviour, but actually practically finding somebody feels a little beyond our scope [35]

- 

###### Relying on other professionals

- tend to rely on support workers to help them socialise but can suggest this as part of the care plan [28]

- However, perhaps social workers / OTs within my team could run groups on safety / appropriate when online dating etc. which would be in keeping with this aim. [31]

###### Relationships are irrelevant for my clients

Desire and capability

- Additionally, the clients have severe mental/physical disabilities and so it is either not something they would desire (e.g. not mentally capable of), or they are too severe to interact with the public/other society members e.g. prone to anger and attacking (e.g. not socially capable of) [5]

Not something service users bring up

- I think that is also how they view me so he wouldn, Äôt necessarily go to me for that (possibly because they don, Äôt deem as appropriate) unless I approached the topic. [5]

##### NEITHER AGREE NOR DISAGREE THEMES

Depends on their level of care: high support not priority

- It depends on the level of care they require, in the more independent housing I would say I would be happy to discuss this but in high support this is not our priority [4]

- Nature of the service/ acute emergency assessments but I would strongly agree for treatment/ community services. who work with people longer term. [P54]

Undecided

- I’m undecided I think it could be good for residents to find meaningful relationships but I wonder if it crosses a boundary to support them into those relationships [20]

#### Question 2, Barriers: ‘How much do the following factors form barriers to “finding a relationship” conversations? - Other barrier (if applicable, please specify)’

##### A relationship is not appropriate for service user

Relationship being detrimental to the service user

- it is probable that a relationship may present another stressor to the service user’s life (moderate amount) [P2]

Service user not capable of relationship

- A view that the individual may not be socially capable of a relationship e.g. if they display traits of aggression [P5]

Service user may put their partner at risk

- The clients who have asked are male and have a history of perpetrators of domestic violence [P7]

- Might be a DV perpetrator themselves so this comes with risks [P25]

Service user not ready for a relationship

- just had a baby, might not be ready yet. [P30]

##### Worrying about not being capable

Liability

- Worry that if it goes wrong will the resident blame me? [P31]

Making things worse

- If a person has a severe learning disability and has always been single, conversations about relationships (if handled incorrectly)may come across as condescending or as if you are making fun of them [P13]

- also concerns that the discussion (where initiated by clinician) stigmatise their single status) [P40]

Being ill equipped

- Also feel inequipped like it would be opening a tin of worms [P44]

Other people are better suited to this work

- I think I’m not necessarily the best person as a consultant psychiatrist carrying out relatively formal reviews - conversations better initiated by people like care coordinators or support workers in a relatively informal setting. [P38]

##### Organisational factors

Not part of routine practice

- Relationship goals not being part of routine assessment [P46]

Colleague hesitation

- Colleagues opinions of whether it’s our role [P49]

Not appropriate context

- starting up relationship problems in a therapeutic session [P2]

Systemic deprioritisation

- Consideration of relationships fits a social/biopsychosocial model of mental health, but many services and service users are still within an ’illness’ model where holistic conversations re: wider relationships may not be a) trained, b) seen as appropriate [P51]

##### Communication issues

Needing therapeutic alliance first

- People not wanting to discuss this until they have built a trusting relationship with the professional [P46]

Service user not asking for support

- The person who we support being able to properly communicate their desire for a relationship even if it is something they might want [P13]

##### External factors

Families being overprotective

- If the family are active in the person we supports lives, they could be overprotective and not want their child to have a relationship e.g. they feel they are too vulnerable or would not want staff to be involved in that way [P13]

- If the family are heavily involved, they can often be overprotective and not want their child to be dating [P5]

Related barriers

- Other support around the person such as family or services being opposed [P46]

- service users’ previous relationship problems [P2]

#### Question 3, Barriers: ‘Which of these barriers do you think is the most significant?’

##### Inappropriate for clinician

Inappropriate in my work role

- feeling it is inappropriate in my work role [P2]

- Worries that is inappropriate and intrusive [P4]

- Concerns re: relevance/appropriateness - not personally, but at service level and service user level. [P51]

Breaking boundaries

- Worries about it breaking professional boundaries [P9]

- Ensuring the support remains boundaries and the service user takes the lead if this is what they wish to do [P10]

- professional boundaries [P17]

- Professional boundaries, [P19]

- The barrier around professional boundaries [P31]

- crossing professional lines [P35]

- Boundaries [P51]

- Boundaries, triggering and intrusiveness [P58]

Intrusive

- Worries that is inappropriate and intrusive [P4]

- worries it might seem too intrusive [P41]

- Being intrusive to the service user. Also, a make worker asking a female might be viewed dimly or inappropriate. [P49]

- Boundaries, triggering and intrusiveness [P58]

Gender issues between client and clinician

- Being intrusive to the service user. Also, a make worker asking a female might be viewed dimly or inappropriate. [P49]

- Nature of the service and being a male member of staff I would not feel comfortable discussing with a female patient and would also be concerned that this would make them uncomfortable. [P54]

- I think gender of staff member [P54]

Relationships not prioritised

- Prioritising other issues [P45]

##### Clinician inability

Lack of training

- Lack of training [P32]

- Lack of management training [P13]

- I have experienced no training or discussions around this therefore naturally you think it might be out of the scope of your professional boundaries. Something about it does feel that way. [P25]

- Lack of training around conversations and how these can be appropriate [P36]

- Lack of training perhaps [P42]

- Lack of training [P47]

Hard to identify clients

- Hard to identify just one and varies from person to person [P38]

Lack of time and resources

- Time [P8]

- Time and appropriate services [P12]

- Time, and feeling unable to help. [P43]

- Time constraints are a factor [P27]

Low clinician confidence

- voulnerability. I’m and experienced nurse although low confidence with this [30]

- not knowing how to support someone to seek a relationship and worry about their vulnerability [P24]

- Time, and feeling unable to help. [P43]

- Not feeling able to help [P57]

- Not being able to do anything to help if they want a relationship [P59]

##### Client factors

Client’s history

- Client’s history of domestic violence [P7]

- Sexual assault [P41]

Client context

- Either there pregnant or have a baby up to the age of 2. May need support for this before getting into a new relationship. [P30]

Vulnerable patients

- vulnerable patients [P19]

- voulnerability. I’m and experienced nurse although low confidence with this [P24]

- not knowing how to support someone to seek a relationship and worry about their vulnerability [P27]

- Patient being vulnerable [P34]

- Sexual assault [P41]

- worries they will be exploited, or exploit someone [P57]

- Boundaries, triggering and intrusiveness [P58]

- worries about service user being vulnerable to exploitation [P62]

##### Organisational factors

Support

- Lack of management support [P5]

- Management support and the general perception of the public about people with mental illness dating [P33]

- Other supports and services seeing this as unimportant or inappropriate [P46]

- Management support [P48]

- Lack of support / training around having these conversations [P56]

- nature of service are largest factors [P54]

Training

- I have experienced no training or discussions around this therefore naturally you think it might be out of the scope of your professional boundaries. Something about it does feel that way. [31]

- Lack of support / training around having these conversations [P56]

##### Societal perception

- Management support and the general perception of the public about people with mental illness dating [P25]

- I would not want to ask someone about finding a relationship if they have not brought it up themselves. This is because I would not want to assume that finding a romantic relationship is something they need or want to do - I don’t want to impose societal expectations of relationships on someone, particularly if struggling with their wellbeing. It would need to be worded more broadly, i.e. are you interested in relationships / is this something that’s important to you? [P51]

##### None

- None, it is part of the role to address this aspect of their lives as well in thinking about the person as a whole [P6]

#### Question 4, Barriers: ‘Is there anything else you’d like to say about these barriers?’

##### More training and support needed

Management and policy

- I think the main barrier is management, it is never really spoken about that we need to help the clients find relationships, especially because there are so many people involved that it would need to be discussed with e.g. the care management, social workers, the family. When we’ve had less severe clients in the past who weee definitely capable of relationships and I think would benefit/enjoy it, it was something that was never spoken about or we were trained in /encouraged to help so it was never something we tried to talk about with them. [17]

- I understand their importance but residents do want someone in their life romantically and I want residents to feel happy it would be helpful if there were more clear guidelines on how we can support a client with this [37]

- Everything you do has to be mostly approved or encouraged by them, so even if you have no personal barriers, without management support, it is not something that can be done. [7]

Staff need training

- more training in this regard [41]

- they are very daughting so would i feel staff would like support arund this [33]

##### Staff may get it wrong

- It could appear as though single staff members are flirting if this conversation was not done effectively [56]

##### Important issue

- it is very important that this topic is spoken about, this is huge in peoples lives [30]

- It seems a shame because romantic relationships are so important and could be the changing factor in someone’s loneliness, suicidality and wellbeing. [31]

- This is a very important area, and I welcome a better understanding of how to navigate these issues and offer a more holistic approach to clients [48]

##### Client needs to be stable before engaging in relationship

- Need to focus on own mental health and wellbeing and care of a baby. Get to know other people but take things slowly if it works out they will understand. having a baby is one of the most difficult things you’ll ever do as a women, and if someone is unwell some awful individuals will take advantage of this. To think about where, when and why your meeting someone new, making friends and family aware so that they know if you do get into any bother. [36]

- Sometimes people, at least in more acute services, might benefit from more stability (e.g. of mpod, of routine, being able to go out) if they are then to find a hood relationship. But this is cery influenced by the fact i wiork in secondary and inpatient services. [51]

#### Question 5, Current practice: ‘Please tell us about any ways in which you, or others you work with, try to help those who express a desire for an intimate / romantic relationship.’

##### Discussions with service user

Just having a conversation

- asking a simple question like “What are your thoughts about romantic relationships?” [P2]

- Just having a conversation about it and finding out their thoughts. [P33]

- conversation, reassurance, practical advice [P35]

- Explore whether they have any pre-existing ideas in mind first and take it from there [P6]

- I ask about satisfaction with intimate relationships as part of an initial screener/putcome measure. [P49]

- conversations re: navigating sex and consent. [P51]

- Exploring ideas around relationships, sex, where they come from. [P56]

Discerning client goals

- to ask whether having a relationship is one of their goals [P40]

- asking what a person would like in the future or work towards [P24]

- I have chats with my clients about what kind of relationship they want. I try to support them by asking how they like to reach out to significant others i.e. in person/online and discuss safety when dating e.g. keeping to public spaces not sharing personal information etc. more of an advisory role as opposed to engaging them directly. [P31]

Discussing barriers

- I guess figure out whats stopping them in the first place or what barriers they have and figure out what we can do to help with those barriers [P41]

Discussing social network

- Discussion of social connections [P4]

- Optimising mental health can sometimes facilitate this, as can helping people to engage in social activities. But this is an indirect effect of something that is done anyway. [P43]

Discussion of sexual needs

- Meeting sexual needs in hospital, social skills, online safety [P48]

##### Teaching and advice

Relationship education

- We try and sensitively explain how consensual relationships work. Remind them how coercive behaviour is wrong and try and guide them on acceptable behaviour [P7]

- explore healthy and unhealthy relationships. To think about the whole picture in keeping themselves safe keep talking about it. [P30]

- I have chats with my clients about what kind of relationship they want. I try to support them by asking how they like to reach out to significant others i.e. in person/online and discuss safety when dating e.g. keeping to public spaces not sharing personal information etc. more of an advisory role as opposed to engaging them directly. [P31]

- We can provide support and encouragement, help people to identify their needs and goals, teach people about communication and relationship skills, and connect people with resources in the community. [P39]

- Talking about and having easy read materials available describing qualities of supportive romantic relationships, and dating safety including online. Role playing starting conversations or using drama or discussion to explore feelings around romance and sexuality. Also sadly safeguarding as sometimes this is expressed in the context of abusive relationships. [P46]

- Meeting sexual needs in hospital, social skills, online safety [P48]

- Discuss safety and healthy relationships and what they look like [P59]

Practical advice

- conversation, reassurance, practical advice [P35]

- I have chats with my clients about what kind of relationship they want. I try to support them by asking how they like to reach out to significant others i.e. in person/online and discuss safety when dating e.g. keeping to public spaces not sharing personal information etc. more of an advisory role as opposed to engaging them directly. [P31]

- I would say they need to get out of their home to try and meet new people and hope they meet someone that way [P50].

- Recommendations re: dating apps, approaches to dating [P51]

##### Active interventions

Increasing access to partners

- Direct them to dating sites o encourage their families/carers to support [P8]

- look into options of where to find a good match - common interests [P21]

- behavioural experiments using dating apps and broadening social opportunities. [P25]

- We can provide support and encouragement, help people to identify their needs and goals, teach people about communication and relationship skills, and connect people with resources in the community. [P39]

- Support to meet people online, discuss progress [P57]

Skills building

- It wouldn’t be about finding a relationship for them but more so about building interpersonal social skills and recognising how they might be vulnerable to exploitation [P10]

- Meeting sexual needs in hospital, social skills, online safety [P48]

Building self-esteem

- Building up self-esteem [P11]

- building self confidence [P51]

Therapeutic interventions

- Motivational interviewing around meeting others [P25]

- Through therapy, clarifying what they want, problrm solving how they might go about it, challenging anxieties or negative thoughts about how it might work out. S [P45]

- In LD service we specifically had a sex and relationships group. It comes up in CAMHS but feels less focused on as a part of the work [P47]

##### None known

Unsure

- I’m not sure! But I’m only at this service once a week so I may miss a lot of these conversations. I have spoken to clients about past relationships, but not about seeking out future ones since no desire was expressed and there were more pressing matters to address. [P32]

Not done in my service

- This is not something that is done in my practice. [P5]

- In 20 years I’ve not witnessed this [P34]

- None [P58]

##### Taking service user’s lead

- If sex or romantic relationships are specifically raised as an identified issue, this will be explored, formulated and any barriers discussed. Intimate/family relationships is a part of the care plan, however it rarely is a priority to focus on within the time frame. [P25]

- Begin to discuss when they bring it up - based around their circumstances there is no guidance [P44]

#### Question 6, Future suggestions: ‘Are there any other ways you think staff in mental health and social care services could support people in finding a relationship (even if these are not current practice in your workplace)?’

##### Opportunities for socialising

Attending social activities outside the service

- Attending group activities in the community, more opportunities to interact with people outside of the service [P4]

- Taking clients to community events e.g. coffee events with other people with disabilities, social dance classes etc. where they can be encouraged to interact with other people in society and potentially form relationships. [P5]

- Unsure, it also feels quite strange to offer a specific intervention around it as you can feel like a dating service and it might feel forced. Where as taking a social intervention and allowing any relationships to come naturally seems the more acceptable way of doing it. [P25]

Facilitation within the service

- Social opportunities in services [P11]

- Possibly some form of safe dating facilitation - many people who use mental health services have relationships with others who do too. Quite a few fears though about the possibility that resulting relationships might turn out to be problematic in some way [P38].

Finding dating services

- Maybe helping with dating sites but again i wouldnt know how to support someone to not get taken advantage of here. [P27]

- I think it would be good to find dating services (free) that is like speed-dating or friendship making. Or provide this within the service or in the wider organisation so that residents can establish positive relationships [P31]

Signposting

- I suppose via signposting to community activities? [P32]

##### More discussion about relationships in service

Asking about barriers to relationships at assessment

- But explicitly asking about thoughts and barriers to finding romantic relationships should be more prominent in assessment questions and interventions around mental health.. [P25]

Group work

- Discussion and group work [P26]

- Offer support in the form of groups to discuss what a healthy relationship looks like.

Do some scenarios and explore what they would do in that situation. [P30]

- Possibly, groups to discuss online dating safety etc. [P43]

Open discussion

- Discussing it more openly and not stigmatising [P33]

- to give permission to patients to state this as a goal and set out how you might support someone in their socialisation to address this [P40]

- Talking about sex and relationships more [P48]

- Open conversion. Looking at roles and routines that might support opportunities. We can explore what the emotional, practical and esteem barriers are. There are always options but it takes the service user a lot to over come this. [P49]

Psychoeducation

- offer some eduation about safety in relaitonships for vulnerable peope including udnerstanding their own comfortable boundaries. [P27]

- Perhaps psycho Ed on healthy relationships, for some people like LD services maybe learning social skills to develop relationships [P47]

- And by helping to psychologically prepare them for a relationship if needed [P32]

##### Indirect support

Signposting to services

- Direct them to relevant services [P8]

- Training! Being aware of organisations to signpost to that set up safe spaces for more vulnerable people to meet someone. [P46]

Skills building

- By working on the individual to build skills mentioned above that’s as far as we should go [P10]

- Maybe some skills support around going on dates e.g conversationa dn social interaction [P27]

- Support with social skills, ensuring the person is being treated for symptoms that may affect confidence etc, or functioning, addressing sexual side effects or symptoms as well. Improving self-esteem, [P42]

##### Systemic change

Needing more knowledge in this area

- more knowledge around this [P21]

- I don’t know, and probably should [P24]

- Training [P34]

- No [P35]

- Unsure [P50]

Organisational change

- improving policies and access to material to meet needs [P48]

##### Should not be part of MHP role

- I do not think it should be part of my role [P7]

### Step 3: Merging seven analyses into four

#### Merge 1: Barriers

##### Provider factors

Provider not feeling able to help

- Worry that if it goes wrong will the resident blame me? [P31]

- If a person has a severe learning disability and has always been single, conversations about relationships (if handled incorrectly)may come across as condescending or as if you are making fun of them [P13]

- also concerns that the discussion (where initiated by clinician) stigmatise their single status) [P40]

- Also feel inequipped like it would be opening a tin of worms [P44]

- I think I’m not necessarily the best person as a consultant psychiatrist carrying out relatively formal reviews - conversations better initiated by people like care coordinators or support workers in a relatively informal setting. [P38]

- voulnerability. I’m and experienced nurse although low confidence with this [P30]

- not knowing how to support someone to seek a relationship and worry about their vulnerability [P24]

- Time, and feeling unable to help. [P43]

- Not feeling able to help [P57]

- Not being able to do anything to help if they want a relationship [P59]

- Hard to identify just one and varies from person to person [P38]

Inappropriate for the provider

- feeling it is inappropriate in my work role [P2]

- Worries that is inappropriate and intrusive [P4]

- Concerns re: relevance/appropriateness - not personally, but at service level and service user level. [P51]

- Worries about it breaking professional boundaries [P9]

- Ensuring the support remains boundaries and the service user takes the lead if this is what they wish to do [P10]

- professional boundaries [P17]

- Professional boundaries, [P19]

- The barrier around professional boundaries [P31]

- crossing professional lines [P35]

- Boundaries [P51]

- Boundaries, triggering and intrusiveness [P58]

- Worries that is inappropriate and intrusive [P4]

- worries it might seem too intrusive [P41]

- Being intrusive to the service user. Also, a make worker asking a female might be viewed dimly or inappropriate. [P49]

- Boundaries, triggering and intrusiveness [P58]

- Being intrusive to the service user. Also, a make worker asking a female might be viewed dimly or inappropriate. [P49]

- Nature of the service and being a male member of staff I would not feel comfortable discussing with a female patient and would also be concerned that this would make them uncomfortable. [P54]

- I think gender of staff member [P54]

- Prioritising other issues [P45]

Communication issues

- The person who we support being able to properly communicate their desire for a relationship even if it is something they might want [P13]

- People not wanting to discuss this until they have built a trusting relationship with the professional [P46]

##### Service user factors

Relationships are inappropriate for the service user

- just had a baby, might not be ready yet. [P30]

- The clients who have asked are male and have a history of perpetrators of domestic violence [P7]

- Might be a DV perpetrator themselves so this comes with risks [P25]

- A view that the individual may not be socially capable of a relationship e.g. if they display traits of aggression [P5]

- it is probable that a relationship may present another stressor to the service user’s life (moderate amount) [P2]

- Client’s history of domestic violence [P7]

- Sexual assault [P41]

- Either there pregnant or have a baby up to the age of 2. May need support for this before getting into a new relationship. [P30]

- vulnerable patients [P19]

- voulnerability. I’m and experienced nurse although low confidence with this [P24]

- not knowing how to support someone to seek a relationship and worry about their vulnerability [P27]

- Patient being vulnerable [P34]

- Sexual assault [P41]

- worries they will be exploited, or exploit someone [P57]

- Boundaries, triggering and intrusiveness [P58]

- worries about service user being vulnerable to exploitation [P62]

- service users’ previous relationship problems [P2]

Service user must be stable

- Need to focus on own mental health and wellbeing and care of a baby. Get to know other people but take things slowly if it works out they will understand. having a baby is one of the most difficult things you’ll ever do as a women, and if someone is unwell some awful individuals will take advantage of this. To think about where, when and why your meeting someone new, making friends and family aware so that they know if you do get into any bother. [36]

- Sometimes people, at least in more acute services, might benefit from more stability (e.g. of mpod, of routine, being able to go out) if they are then to find a hood relationship. But this is cery influenced by the fact i wiork in secondary and inpatient services. [51]

##### Organisational factors

Lack of support

- Colleagues opinions of whether it’s our role [P49]

- Consideration of relationships fits a social/biopsychosocial model of mental health, but many services and service users are still within an ’illness’ model where holistic conversations re: wider relationships may not be a) trained, b) seen as appropriate [P51]

- Lack of management support [P5]

- Management support and the general perception of the public about people with mental illness dating [P33]

- Other supports and services seeing this as unimportant or inappropriate [P46]

- Management support [P48]

- Lack of support / training around having these conversations [P56]

- nature of service are largest factors [P54]

- I think the main barrier is management, it is never really spoken about that we need to help the clients find relationships, especially because there are so many people involved that it would need to be discussed with e.g. the care management, social workers, the family. When we’ve had less severe clients in the past who weee definitely capable of relationships and I think would benefit/enjoy it, it was something that was never spoken about or we were trained in /encouraged to help so it was never something we tried to talk about with them. [17]

- Everything you do has to be mostly approved or encouraged by them, so even if you have no personal barriers, without management support, it is not something that can be done. [7]

Lack of training

- I have experienced no training or discussions around this therefore naturally you think it might be out of the scope of your professional boundaries. Something about it does feel that way. [31]

- Lack of support / training around having these conversations [P56]

- Lack of training [P32]

- Lack of management training [P13]

- Lack of training around conversations and how these can be appropriate [P36]

- Lack of training perhaps [P42]

- Lack of training [P47]

- more training in this regard [41]

- they are very daughting so would i feel staff would like support arund this [33]

- It could appear as though single staff members are flirting if this conversation was not done effectively [56]

Lack of policy

- Relationship goals not being part of routine assessment [P46]

- starting up relationship problems in a therapeutic session [P2]

- I understand their importance but residents do want someone in their life romantically and I want residents to feel happy it would be helpful if there were more clear guidelines on how we can support a client with this [37]

Lack of time and resources

- Time [P8]

- Time and appropriate services [P12]

- Time, and feeling unable to help. [P43]

- Time constraints are a factor [P27]

- Even though this is important, it is difficult to address these issues in the time slot given to me to see the service user. I also do not have access to resources or skills to be able to help someone like that. [8]

- Also, there is barely enough time to do the core aspects of my job, so in my opinion there is very unlikely to be resource for this. [31]

- services are usually very limited in the type of therapeutic intervention we are ’allowed’ to offer due to funding and commissioning, and focused on particular mental-health related outcomes (which while linked to relationships, are not usually defined as such). [P51]

- 

##### External factors

Families being overprotective

- If the family are active in the person we supports lives, they could be overprotective and not want their child to have a relationship e.g. they feel they are too vulnerable or would not want staff to be involved in that way [P13]

- If the family are heavily involved, they can often be overprotective and not want their child to be dating [P5]

Societal perception

- Other support around the person such as family or services being opposed [P46]

- Management support and the general perception of the public about people with mental illness dating [P25]

- I would not want to ask someone about finding a relationship if they have not brought it up themselves. This is because I would not want to assume that finding a romantic relationship is something they need or want to do - I don’t want to impose societal expectations of relationships on someone, particularly if struggling with their wellbeing. It would need to be worded more broadly, i.e. are you interested in relationships / is this something that’s important to you? [P51]

##### None

- None, it is part of the role to address this aspect of their lives as well in thinking about the person as a whole [P6]

#### Merge 2: Current practice and suggestions

##### Current practice

Preparing the service user for the dating world

- It wouldn’t be about finding a relationship for them but more so about building interpersonal social skills and recognising how they might be vulnerable to exploitation [P10]

- Meeting sexual needs in hospital, social skills, online safety [P48]

- Building up self-esteem [P11]

- building self confidence [P51]

- Motivational interviewing around meeting others [P25]

- Through therapy, clarifying what they want, problrm solving how they might go about it, challenging anxieties or negative thoughts about how it might work out. S [P45]

- In LD service we specifically had a sex and relationships group. It comes up in CAMHS but feels less focused on as a part of the work [P47]

- We try and sensitively explain how consensual relationships work. Remind them how coercive behaviour is wrong and try and guide them on acceptable behaviour [P7]

- explore healthy and unhealthy relationships. To think about the whole picture in keeping themselves safe keep talking about it. [P30]

- I have chats with my clients about what kind of relationship they want. I try to support them by asking how they like to reach out to significant others i.e. in person/online and discuss safety when dating e.g. keeping to public spaces not sharing personal information etc. more of an advisory role as opposed to engaging them directly. [P31]

- We can provide support and encouragement, help people to identify their needs and goals, teach people about communication and relationship skills, and connect people with resources in the community. [P39]

- Talking about and having easy read materials available describing qualities of supportive romantic relationships, and dating safety including online. Role playing starting conversations or using drama or discussion to explore feelings around romance and sexuality. Also sadly safeguarding as sometimes this is expressed in the context of abusive relationships. [P46]

- Meeting sexual needs in hospital, social skills, online safety [P48]

- Discuss safety and healthy relationships and what they look like [P59]

- conversation, reassurance, practical advice [P35]

- I have chats with my clients about what kind of relationship they want. I try to support them by asking how they like to reach out to significant others i.e. in person/online and discuss safety when dating e.g. keeping to public spaces not sharing personal information etc. more of an advisory role as opposed to engaging them directly. [P31]

- I would say they need to get out of their home to try and meet new people and hope they meet someone that way [P50].

- Recommendations re: dating apps, approaches to dating [P51]

Discussions with service user about relationships

- asking a simple question like “What are your thoughts about romantic relationships?” [P2]

- Just having a conversation about it and finding out their thoughts. [P33]

- conversation, reassurance, practical advice [P35]

- Explore whether they have any pre-existing ideas in mind first and take it from there [P6]

- I ask about satisfaction with intimate relationships as part of an initial screener/putcome measure. [P49]

- conversations re: navigating sex and consent. [P51]

- Exploring ideas around relationships, sex, where they come from. [P56]

- to ask whether having a relationship is one of their goals [P40]

- asking what a person would like in the future or work towards [P24]

- I have chats with my clients about what kind of relationship they want. I try to support them by asking how they like to reach out to significant others i.e. in person/online and discuss safety when dating e.g. keeping to public spaces not sharing personal information etc. more of an advisory role as opposed to engaging them directly. [P31]

- I guess figure out whats stopping them in the first place or what barriers they have and figure out what we can do to help with those barriers [P41]

- Discussion of social connections [P4]

- Optimising mental health can sometimes facilitate this, as can helping people to engage in social activities. But this is an indirect effect of something that is done anyway. [P43]

- Meeting sexual needs in hospital, social skills, online safety [P48]

Increasing access to partners

- Direct them to dating sites o encourage their families/carers to support [P8]

- look into options of where to find a good match - common interests [P21]

- behavioural experiments using dating apps and broadening social opportunities. [P25]

- We can provide support and encouragement, help people to identify their needs and goals, teach people about communication and relationship skills, and connect people with resources in the community. [P39]

- Support to meet people online, discuss progress [P57]

##### Suggestions for future practice

Psycho-education and skills work

- Discussion and group work [P26]

- Offer support in the form of groups to discuss what a healthy relationship looks like.

Do some scenarios and explore what they would do in that situation. [P30]

- Possibly, groups to discuss online dating safety etc. [P43]

- offer some eduation about safety in relaitonships for vulnerable peope including udnerstanding their own comfortable boundaries. [P27]

- Perhaps psycho Ed on healthy relationships, for some people like LD services maybe learning social skills to develop relationships [P47]

- And by helping to psychologically prepare them for a relationship if needed [P32]

- By working on the individual to build skills mentioned above that’s as far as we should go [P10]

- Maybe some skills support around going on dates e.g conversationa dn social interaction [P27]

- Support with social skills, ensuring the person is being treated for symptoms that may affect confidence etc, or functioning, addressing sexual side effects or symptoms as well. Improving self-esteem, [P42]

Open discussion about relationships in service

- But explicitly asking about thoughts and barriers to finding romantic relationships should be more prominent in assessment questions and interventions around mental health.. [P25]

- Discussing it more openly and not stigmatising [P33]

- to give permission to patients to state this as a goal and set out how you might support someone in their socialisation to address this [P40]

- Talking about sex and relationships more [P48]

- Open conversion. Looking at roles and routines that might support opportunities. We can explore what the emotional, practical and esteem barriers are. There are always options but it takes the service user a lot to over come this. [P49]

Increasing access to partners

- Attending group activities in the community, more opportunities to interact with people outside of the service [P4]

- Taking clients to community events e.g. coffee events with other people with disabilities, social dance classes etc. where they can be encouraged to interact with other people in society and potentially form relationships. [P5]

- Unsure, it also feels quite strange to offer a specific intervention around it as you can feel like a dating service and it might feel forced. Where as taking a social intervention and allowing any relationships to come naturally seems the more acceptable way of doing it. [P25]

- Social opportunities in services [P11]

- Possibly some form of safe dating facilitation - many people who use mental health services have relationships with others who do too. Quite a few fears though about the possibility that resulting relationships might turn out to be problematic in some way [P38].

- Maybe helping with dating sites but again i wouldnt know how to support someone to not get taken advantage of here. [P27]

- I think it would be good to find dating services (free) that is like speed-dating or friendship making. Or provide this within the service or in the wider organisation so that residents can establish positive relationships [P31]

Systemic change

- more knowledge around this [P21]

- I don’t know, and probably should [P24]

- Training [P34]

- No [P35]

- Unsure [P50]

- improving policies and access to material to meet needs [P48]

Signposting

- Direct them to relevant services [P8]

- Training! Being aware of organisations to signpost to that set up safe spaces for more vulnerable people to meet someone. [P46]

- I suppose via signposting to community activities? [P32]

## Appendix C Collated quantitative results tables

**Table 1.**
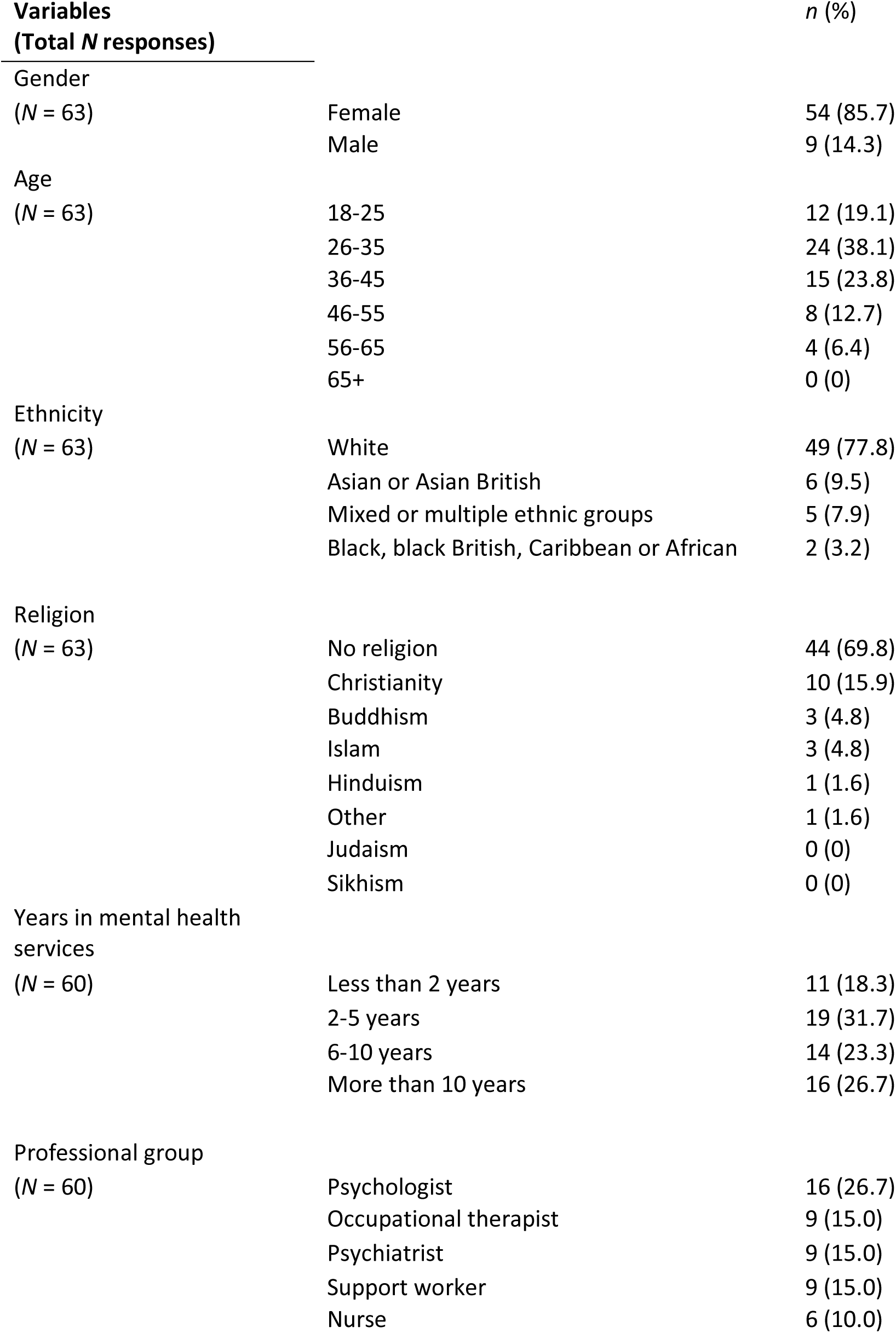
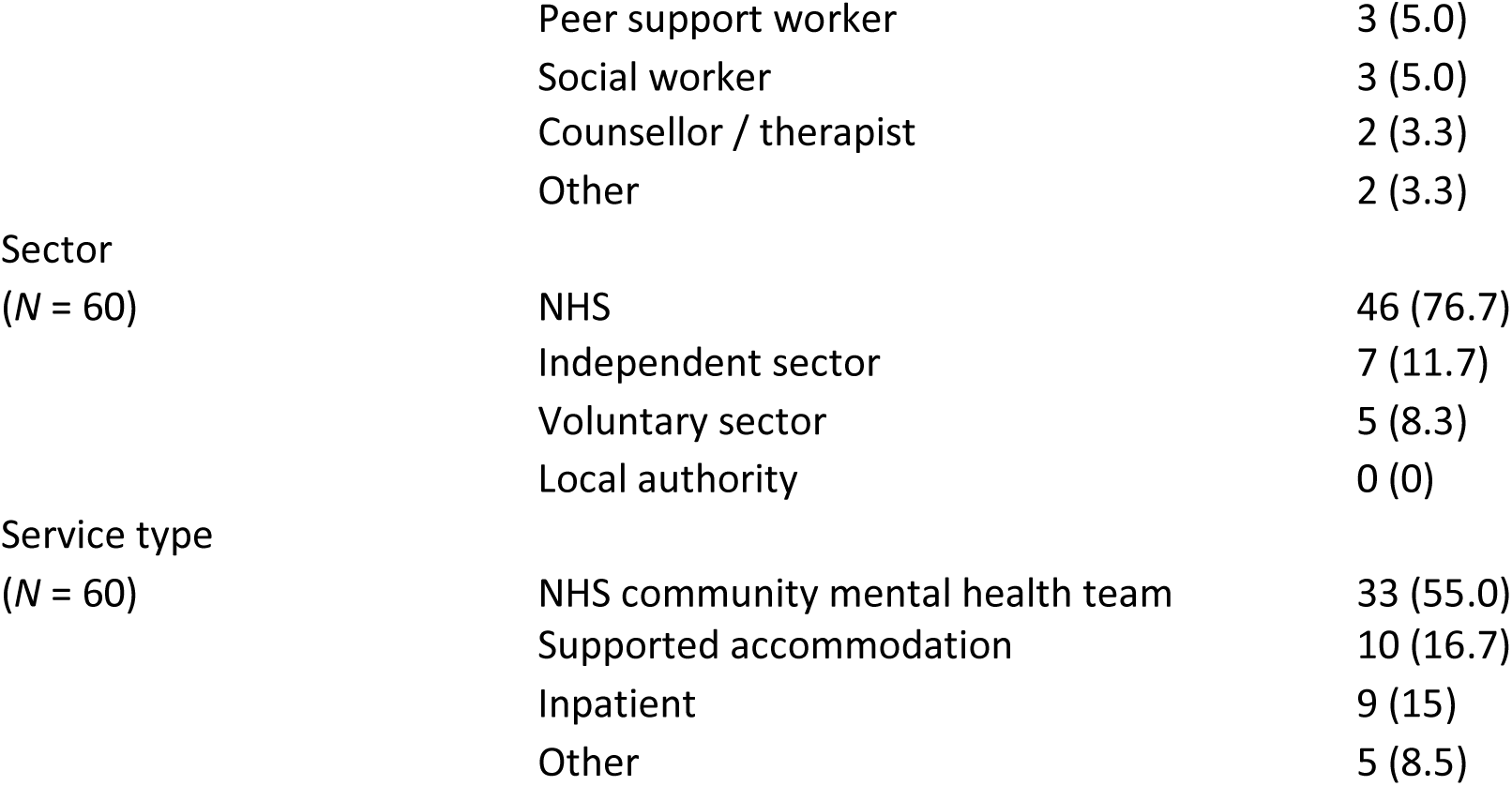
Participant characteristics

**Table 2.** Ratings of agreement as to the appropriateness of ‘finding a relationship’ conversations

| Agreement as to appropriateness of 'finding a relationship' conversations | ***n* (%)**<br>(total N = 50) |
| --- | --- |
| Strongly agree | 10 (20.0) |
| Somewhat agree | 25 (50.0) |
| Somewhat disagree | 11 (22.0) |
| Strongly disagree | 4 (8.0) |

**Table 4.**
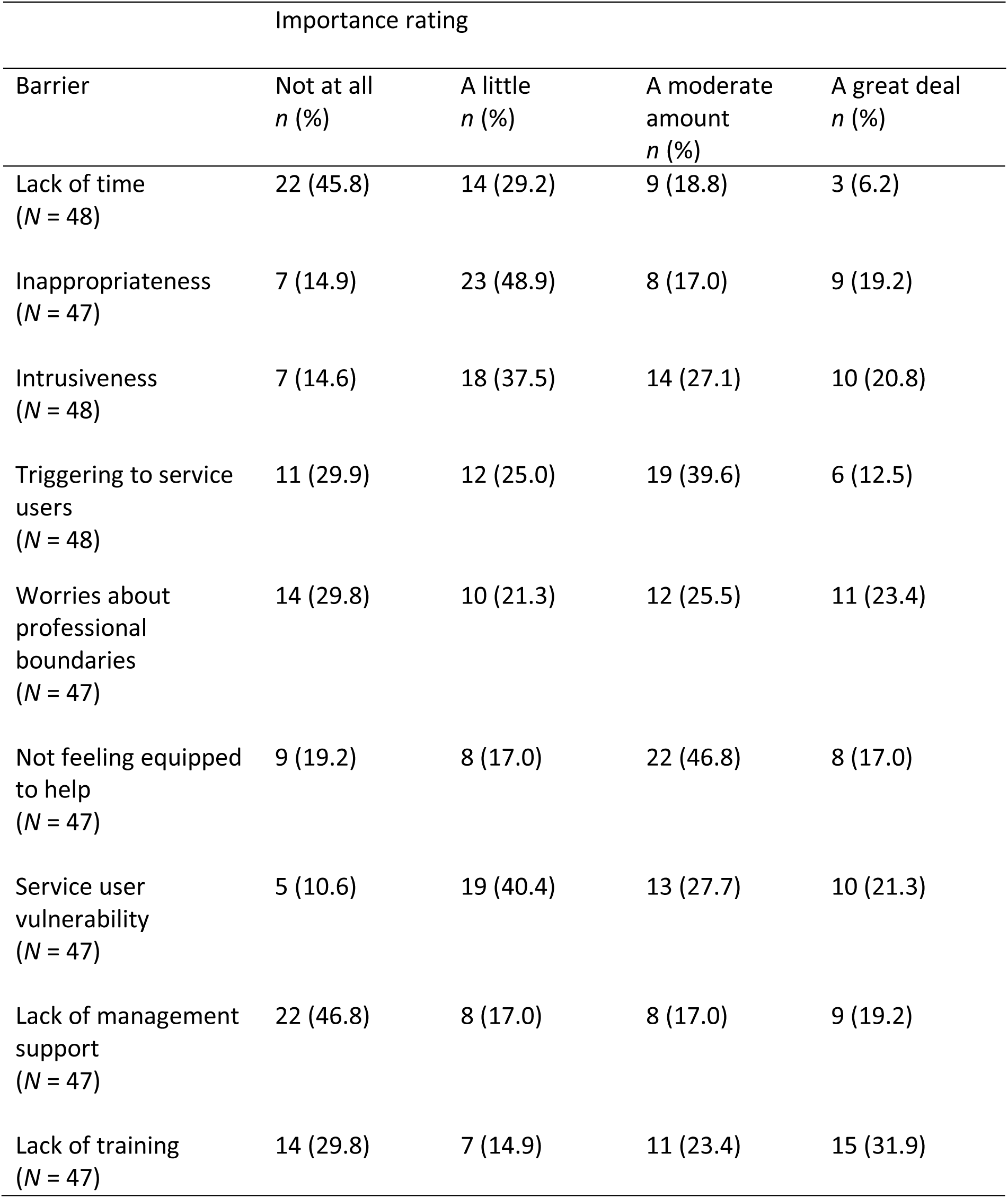
Participant ratings of the importance of potential barriers to helping service users find intimate relationships.

**Table 6.**
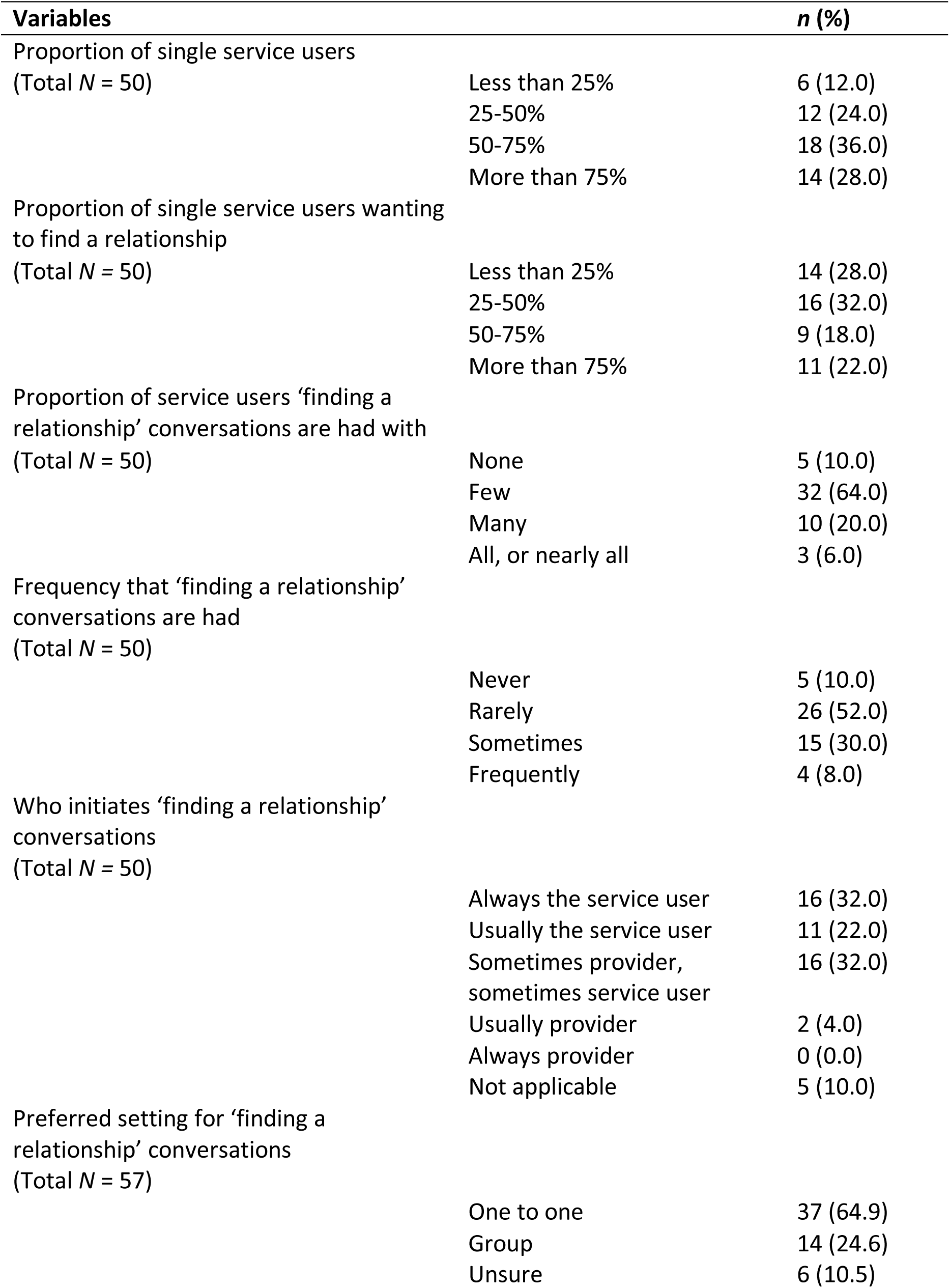
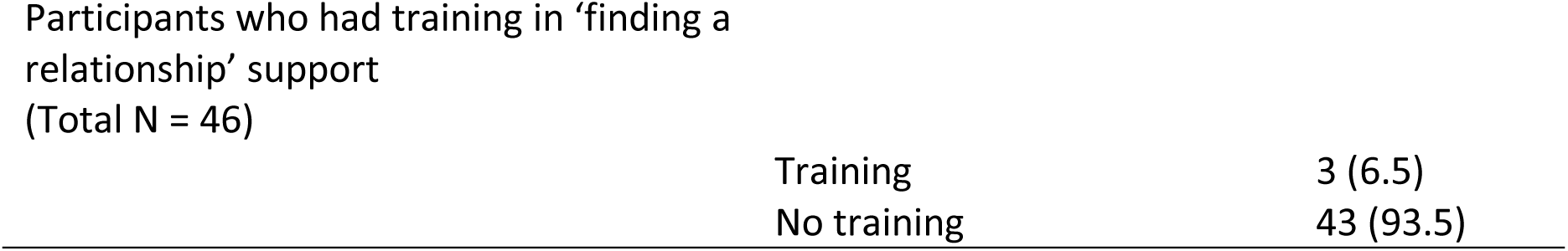
Quantitative findings regarding the nature of ‘finding a relationship’ conversations

## References

1. World Health Organization. Defining sexual health: report of a technical consultation on sexual health, 28-31 January 2002, Geneva: World Health Organization; 2006.

2. Gascoyne S, Hughes E, McCann E, Quinn C. The sexual health and relationship needs of people with severe mental illness. Journal of psychiatric and mental health nursing. 2016;23(5):338–43.

3. McCann E, Donohue G, de Jager J, Nugter A, Stewart J, Eustace-Cook J. Sexuality and intimacy among people with serious mental illness: a qualitative systematic review. JBI Evidence Synthesis. 2019;17(1):74–125.

4. Furman W, Hand LS. The slippery nature of romantic relationships: Issues in definition and differentiation. Romance and sex in adolescence and emerging adulthood: Risks and opportunities. 2006;11:171–8.

5. Diener E, Seligman ME. Very happy people. Psychological science. 2002;13(1):81–4.

6. Kawachi I, Berkman LF. Social ties and mental health. Journal of Urban health. 2001;78:458–67.

7. Taylor ZE, Doane LD, Eisenberg N. Transitioning from high school to college: Relations of social support, ego-resiliency, and maladjustment during emerging adulthood. Emerging Adulthood. 2014;2(2):105–15.

8. Kansky J. What’s love got to do with it? Romantic relationships and well-being. Handbook of well-being. 2018:1–24.

9. Ecker J, Cherner R, Rae J, Czechowski K. Sexual intimacy, mental illness, and homelessness. American Journal of Community Psychology. 2018;61(1-2):131–40.

10. Stevenson C, Neale J. ‘We did more rough sleeping just to be together’–Homeless drug users’ romantic relationships in hostel accommodation. Drugs: Education, Prevention and Policy. 2012;19(3):234–43.

11. Brown RA, Kennedy DP, Tucker JS, Golinelli D, Wenzel SL. Monogamy on the street: A mixed methods study of homeless men. Journal of mixed methods research. 2013;7(4):328–46.

12. Rayburn RL, Corzine J. Your shelter or mine? Romantic relationships among the homeless. Deviant Behavior. 2010;31(8):756–74.

13. Ryan GW, Stern SA, Hilton L, Tucker JS, Kennedy DP, Golinelli D, et al. When, where, why and with whom homeless women engage in risky sexual behaviors: A framework for understanding complex and varied decision-making processes. Sex Roles. 2009;61:536–53.

14. Wang J, Mann F, Lloyd-Evans B, Ma R, Johnson S. Associations between loneliness and perceived social support and outcomes of mental health problems: a systematic review. BMC Psychiatry. 2018;18(1):156.

15. Victor CR, Yang K. The prevalence of loneliness among adults: a case study of the United Kingdom. The Journal of psychology. 2012;146(1-2):85–104.

16. Meltzer H, Bebbington P, Dennis MS, Jenkins R, McManus S, Brugha TS. Feelings of loneliness among adults with mental disorder. Social psychiatry and psychiatric epidemiology. 2013;48:5–13.

17. Cacioppo S, Grippo AJ, London S, Goossens L, Cacioppo JT. Loneliness:Clinical Import and Interventions. Perspectives on Psychological Science. 2015;10(2):238–49.

18. Weiss R. Loneliness: The experience of emotional and social isolation: MIT press; 1975.

19. Hawkley LC, Browne MW, Cacioppo JT. How can I connect with thee? Let me count the ways. Psychological Science. 2005;16(10):798–804.

20. Cacioppo JT, Patrick W. Loneliness: Human nature and the need for social connection: WW Norton & Company; 2008.

21. Hughes ME, Waite LJ, Hawkley LC, Cacioppo JT. A short scale for measuring loneliness in large surveys: Results from two population-based studies. Research on aging. 2004;26(6):655–72.

22. Ma R, Mann F, Wang J, Lloyd-Evans B, Terhune J, Al-Shihabi A, et al. The effectiveness of interventions for reducing subjective and objective social isolation among people with mental health problems: a systematic review. Social Psychiatry and Psychiatric Epidemiology. 2020;55(7):839–76.

23. Berger-Merom R, Zisman-Ilani Y, Jones N, Roe D. Addressing sexuality and intimate relations in community mental health services for people with serious mental illness: A qualitative study of mental health practitioners’ experiences. Psychiatric Rehabilitation Journal. 2022;45(2):170.

24. Tennille J, Bohrman C, Barrenger S, Compton E, Meduna E, Klein L. Behavioral Health Provider Attitudes and Beliefs about Sexuality and Intimacy: Findings from a Mixed Method Design. Community Mental Health Journal. 2022;58(3):444–53.

25. Chuah FLH, Haldane VE, Cervero-Liceras F, Ong SE, Sigfrid LA, Murphy G, et al. Interventions and approaches to integrating HIV and mental health services: a systematic review. Health policy and planning. 2017;32(suppl_4):iv27–iv47.

26. Ford JV, Barnes R, Rompalo A, Hook III EW. Sexual health training and education in the US. Public Health Reports. 2013;128(2_suppl1):96–101.

27. Pandor A, Kaltenthaler E, Higgins A, Lorimer K, Smith S, Wylie K, et al. Sexual health risk reduction interventions for people with severe mental illness: a systematic review. BMC public health. 2015;15:1–13.

28. Gómez-López M, Viejo C, Ortega-Ruiz R. Well-being and romantic relationships: A systematic review in adolescence and emerging adulthood. International journal of environmental research and public health. 2019;16(13):2415.

29. de Jager J, Cirakoglu B, Nugter A, van Os J. Intimacy and its barriers: A qualitative exploration of intimacy and related struggles among people diagnosed with psychosis. Psychosis. 2017;9(4):301–9.

30. Boucher M-E, Groleau D, Whitley R. Recovery and severe mental illness: The role of romantic relationships, intimacy, and sexuality. Psychiatric Rehabilitation Journal. 2016;39(2):180.

31. Perry BL, Wright ER. The sexual partnerships of people with serious mental illness. Journal of Sex Research. 2006;43(2):174–81.

32. Mizock L, La Mar K, DeMartini L, Stringer J. Relational resilience: intimate and romantic relationship experiences of women with serious mental illness. Journal of Relationships Research. 2019;10:e5.

33. Corrigan P. How stigma interferes with mental health care. American psychologist. 2004;59(7):614.

34. Jorm AF, Oh E. Desire for social distance from people with mental disorders. Australian & New Zealand Journal of Psychiatry. 2009;43(3):183–200.

35. Boysen GA. Stigma Toward People with Mental Illness as Potential Sexual and Romantic Partners. Evolutionary Psychological Science. 2017;3(3):212–23.

36. Corrigan PW, Watson AC. The paradox of self-stigma and mental illness. Clinical psychology: Science and practice. 2002;9(1):35.

37. Wainberg ML, Cournos F, Wall MM, Norcini Pala A, Mann CG, Pinto D, et al. Mental illness sexual stigma: Implications for health and recovery. Psychiatric rehabilitation journal. 2016;39(2):90.

38. Lasalvia A, Zoppei S, Van Bortel T, Bonetto C, Cristofalo D, Wahlbeck K, et al. Global pattern of experienced and anticipated discrimination reported by people with major depressive disorder: a cross-sectional survey. The Lancet. 2013;381(9860):55-62.

39. Perlman CM, Martin L, Hirdes JP, Curtin-Telegdi N, Pérez E, Rabinowitz T. Prevalence and Predictors of Sexual Dysfunction in Psychiatric Inpatients. Psychosomatics. 2007;48(4):309–18.

40. Eager S, Killaspy H, Mezey G, Downey M, Lloyd-Evans B. Understanding the social inclusion needs of people living in mental health supported accommodation. medRxiv. 2023:2023.05. 04.23289515.

41. Mazza M, Marano G, Del Castillo AG, Chieffo D, Monti L, Janiri D, et al. Intimate partner violence: A loop of abuse, depression and victimization. World journal of psychiatry. 2021;11(6):215.

42. White R, Haddock G, Varese F, Haarmans M. “Sex isn’t everything”: views of people with experience of psychosis on intimate relationships and implications for mental health services. BMC psychiatry. 2021;21(1):307.

43. Jacob K. Recovery model of mental illness: A complementary approach to psychiatric care. SAGE Publications Sage India: New Delhi, India; 2015. p. 117–9.

44. Barnes D, Boland B, Linhart K, Wilson K. Personalisation and social care assessment–the Care Act 2014. BJPsych Bulletin. 2017;41(3):176–80.

45. White R, Haddock G, Varese F. Supporting the intimate relationship needs of service users with psychosis: What are the barriers and facilitators? Journal of Mental Health. 2019.

46. Dyer K, das Nair R. Why don’t healthcare professionals talk about sex? A systematic review of recent qualitative studies conducted in the United kingdom. J Sex Med. 2013;10(11):2658–70.

47. Hughes E, Edmondson AJ, Onyekwe I, Quinn C, Nolan F. Identifying and addressing sexual health in serious mental illness: views of mental health staff working in two National Health Service organizations in England. International Journal of Mental Health Nursing. 2018;27(3):966–74.

48. Tennille J, Solomon P, Blank M. Case managers discovering what recovery means through an HIV prevention intervention. Community mental health journal. 2010;46:486–93.

49. Miller SA, Byers ES. Practicing Psychologists’ Sexual Intervention Self-Efficacy and Willingness to Treat Sexual Issues. Archives of Sexual Behavior. 2012;41(4):1041–50.

50. Urry K, Chur-Hansen A, Khaw C. ‘It’s just a peripheral issue’: A qualitative analysis of mental health clinicians’ accounts of (not) addressing sexuality in their work. International Journal of Mental Health Nursing. 2019;28(6):1278–87.

51. Hendry A, Snowden A, Brown M. When holistic care is not holistic enough: The role of sexual health in mental health settings. Journal of clinical nursing. 2018;27(5-6):1015–27.

52. Reissing ED, Giulio GD. Practicing clinical psychologists’ provision of sexual health care services. Professional Psychology: Research and Practice. 2010;41(1):57.

53. Östman M. Low satisfaction with sex life among people with severe mental illness living in a community. Psychiatry Research. 2014;216(3):340–5.

54. Howden M, Zipple AM, Tyrrell WF. Dating skills for residential consumers. Psychosocial Rehabilitation Journal. 1994;18(2):67.

55. Hache-Labelle C, Abdel-Baki A, Lepage M, Laurin AS, Guillou A, Francoeur A, et al. Romantic relationship group intervention for men with early psychosis: A feasibility, acceptability and potential impact pilot study. Early Intervention in Psychiatry. 2021;15(4):753–61.

56. Dubreucq M, Lysaker PH, Dubreucq J. A qualitative exploration of stakeholders’ perspectives on the experiences, challenges, and needs of persons with serious mental illness as they consider finding a partner or becoming parent. Front Psychiatry. 2022;13:1066309.

57. Braun V, Clarke V. Reflecting on reflexive thematic analysis. Qualitative research in sport, exercise and health. 2019;11(4):589–97.

58. Braun V, Clarke V. Toward good practice in thematic analysis: Avoiding common problems and be (com) ing a knowing researcher. International journal of transgender health. 2023;24(1):1–6.

59. Forrester-Jones R, Dixon J, Jaynes B. Exploring romantic need as part of mental health social care practice. Disability & Society. 2023:1–23.

60. Clarke V, Braun V, Hayfield N. Thematic analysis. Qualitative psychology: A practical guide to research methods. 2015;3:222–48.

61. Fereday J, Muir-Cochrane E. Demonstrating rigor using thematic analysis: A hybrid approach of inductive and deductive coding and theme development. International journal of qualitative methods. 2006;5(1):80–92.

62. Gutheil TG, Gabbard GO. The concept of boundaries in clinical practice: theoretical and risk- management dimensions. The American journal of psychiatry. 1993;150(2):188–96.

63. Zur O. Power in psychotherapy and counseling. The Zur Institute. 2014.

64. Pope KS, Keith-Spiegel P. A practical approach to boundaries in psychotherapy: Making decisions, bypassing blunders, and mending fences. Journal of clinical psychology. 2008;64(5):638–52.

65. Dobal MT, Torkelson DJ. Making decisions about sexual rights in psychiatric facilities. Archives of Psychiatric Nursing. 2004;18(2):68–74.

66. Mike Slade, Ph.D. Everyday Solutions for Everyday Problems: How Mental Health Systems Can Support Recovery. Psychiatric Services. 2012;63(7):702–4.

67. Barnett P, Steare T, Dedat Z, Pilling S, McCrone P, Knapp M, et al. Interventions to improve social circumstances of people with mental health conditions: a rapid evidence synthesis. BMC Psychiatry. 2022;22(1):302.

68. Robbins PC, Callahan L, Monahan J. Perceived coercion to treatment and housing satisfaction in housing-first and supportive housing programs. Psychiatric services. 2009;60(9):1251–3.

69. Pogoda TK, Cramer IE, Rosenheck RA, Resnick SG. Qualitative analysis of barriers to implementation of supported employment in the Department of Veterans Affairs. Psychiatric Services. 2011;62(11):1289–95.

70. Virtzberg-Rofè D, Rofè T, Rudnick A. Free online dating site for people with mental illness. Psychiatric Services. 2014;65(9):1178-.

71. Claiborne N, Rizzo VM. Addressing sexual issues in individuals with chronic health conditions. Health & Social Work. 2006;31(3):221.

72. Taylor B, Davis S. The extended PLISSIT model for addressing the sexual wellbeing of individuals with an acquired disability or chronic illness. Sexuality and Disability. 2007;25:135–9.

73. Mick JM. Using the BETTER model to assess sexuality. Number 1/February 2004. 2007;8(1):84-6.

74. Khakbazan Z, Daneshfar F, Behboodi-Moghadam Z, Nabavi SM, Ghasemzadeh S, Mehran A. The effectiveness of the Permission, Limited Information, Specific suggestions, Intensive Therapy (PLISSIT) model based sexual counseling on the sexual function of women with Multiple Sclerosis who are sexually active. Multiple sclerosis and related disorders. 2016;8:113–9.

75. Morison LT, C; John, M. Are mental health services inherently feminised? BPS BPS; 2014 [Available from: https://www.bps.org.uk/psychologist/are-mental-health-services-inherently-feminised.]

76. Bates C. Supported Loving–developing a national network to support positive intimate relationships for people with learning disabilities. Tizard Learning Disability Review. 2019;24(1):13–9.

